# Does Intermittent Nutrition Enterally Normalise hormonal and metabolic responses to feeding in critically ill adults? A protocol for the DINE-Normal proof-of-concept randomised parallel group study

**DOI:** 10.1101/2024.03.14.24303591

**Authors:** Clodagh E Beattie, Matt Thomas, Borislava Borislavova, Harry A Smith, Michael Ambler, Paul White, Kati Hayes, Danielle Milne, Aravind V Ramesh, Javier T Gonzalez, James A Betts, Anthony E Pickering

## Abstract

**Introduction:** Over half of patients who spend >48 hours in the intensive care unit (ICU) are fed via a nasogastric (NG) tube. Current guidance recommends continuous delivery of feed throughout the day and night. Emerging evidence from healthy human studies shows that NG feeding in an intermittent pattern (rather than continuous) promotes phasic hormonal, digestive and metabolic responses that are important for effective nutrition. It is not yet known whether this will translate to the critically ill population. Here we present the protocol for a proof-of-concept study comparing diurnal intermittent versus continuous feeding for patients in the intensive care unit.

**Methods and Analysis:** The study is a single-centre, prospective, randomised, open-label trial comparing intermittent enteral nutrition with the current standard practice of continuous enteral feeding. It aims to recruit participants (n=30) needing enteral nutrition via an NG tube for >24 hours who will be randomised to a diurnal intermittent or a continuous feeding regime with equivalent nutritional value. The primary outcome is peak plasma insulin / c-peptide within 3 hours of delivering the morning bolus feed on the second study day, compared to that seen in the continuous feed delivery group at the same timepoint. Secondary outcomes include feasibility, tolerability, efficacy and metabolic / hormonal profiles.

**Ethics and Dissemination:** This trial has been registered prospectively with the Clinical Trials Registry (clinicaltrials.gov NCT06115044). We obtained ethical approval from the Wales Research Ethics Committee 3 prior to data collection (reference 23/WA/0297). We will publish the results of this study in an open-access peer-reviewed journal.

**Strengths and Limitations of this Study:**

- Having studied metabolic consequences of intermittent versus continuous nasogastric feed delivery in healthy volunteers, our trial team is well positioned to explore the direct ‘bench-to-bedside’ exploration of the interaction between critical illness and the mode of feed delivery through detailed profiling of metabolic hormones.
- The intermittent feeding intervention in this study has been carefully considered to replicate diurnal rhythms, with sufficient fasting between feeding cycles, to allow us to capture a difference in response to continuous feeding if one exists.
- The findings of this study will inform the feasibility and design of a trial of the effectiveness of intermittent diurnal nutrition in critically ill adults.
- This is an open-label, single-centre study, which may bias the interpretation and limit the generalisability of the results.

## INTRODUCTION

In the UK around 200,000 patients are admitted to critical care units annually (icnarc.org), and between 30-50% of those patients are malnourished at the time of admission.^1^ International guidelines emphasise the importance of providing early adequate enteral nutrition (EN) for critically ill patients.^2–4^ Approximately half of these critically ill patients will be fed via a nasogastric (NG) tube because they are unable to feed themselves for a prolonged period.^5^ There is uncertainty about the optimal NG feeding regimen in these critically ill patients and nutritional targets are frequently missed.^6^

The current standard of care is continuous delivery of feed, throughout the day and night. This feeding pattern is unphysiological, both in the sense that it fails to trigger acute mealtime metabolic/hormonal and gastrointestinal responses and that there are none of the usual post-prandial periods aligned with circadian rhythms in metabolism. Other feed patterns have been described, such as intermittent feeding, where feed is delivered in divided doses over a period of between 20 minutes to an hour, with breaks of several hours in between. Bolus feeding describes a similar pattern, but with shorter duration of feeds (generally 5-10 minutes). The terms intermittent and bolus feeding are often used interchangeably in the literature.^7^ The existent recommendation in favour of continuous NG feeding is based on the results of meta-analyses suggesting intermittent administration increases the risk of adverse gastrointestinal sequalae including: diarrhoea, vomiting, constipation, abdominal distension and aspiration.^4,8^ However, the evidence synthesised in these meta-analyses was deemed to be of low quality, with small numbers of participants.

Several studies of intermittent feeding have been conducted in the Intensive Care Unit (ICU). These have been relatively small, each recruiting fewer than 200 patients, and have failed to demonstrate improvements in morbidity or mortality.^7,9,10^ These intermittent feeding regimens frequently continue to deliver food during the night or do not include a prolonged fasting period which may diminish the potential benefits.^10–16^ Primary outcomes have focused on measures of gastrointestinal tolerance while largely neglecting potentially important hormonal, metabolic, circadian, sleep or delirium outcomes.^7,9,17^ Only a few of these studies have published mortality and length of stay data as secondary outcomes, and none have assessed longer-term functional patient-centred outcomes. A recent review of the research agenda for nutrition in the ICU cited the need for phase II trials to study the effects of different feeding patterns on biological markers of metabolism with a view to a phase III RCT focusing on mortality and physical function as the main outcomes.^2^

Diurnal feeding describes the alignment of wake/light cycles with feeding and sleep/dark cycles with fasting. There are several reasons why diurnal intermittent feeding might be beneficial for critically ill patients, compared to the usual continuous administration of NG feed. Of particular interest are the potential metabolic benefits of intermittent diurnal feeding. Evidence from animal studies highlights the importance of circadian rhythms in regulating digestion and metabolism.^18–20^ Hormonal secretion, regulated by the circadian clock, affects the capacity for glucose, protein, and lipid metabolism.^7,21^ Critically ill patients are particularly susceptible to dysregulation of the circadian rhythm, which is influenced heavily by exogeneous cues (zeitgebers) such as the timing of nutritional intake.^22,23^

Studies in humans have shown that the patterns of feeding/fasting cycles affect metabolism. Muscle protein synthesis - of great importance in the critically ill population - is stimulated more effectively by ‘pulsed’ ingestion of protein than a continuous supply of amino acids. This is thought to be due to the ‘leucine trigger hypothesis’, where a critical dose of protein is required to maximise anabolism.^24–26^ In health, with normal oral intake of meals, metabolic hormones such as insulin and ghrelin are released in a pulsatile manner in response to feeding.^27^ This pattern is maintained with intermittent NG feeding in healthy adults, but lost with continuous feeding, with important implications for skeletal muscle autophagy and gut motility.^28^ Intermittent feeding was also shown to increase splanchnic blood flow, which may be beneficial for gastrointestinal tolerance of enteral nutrition.^28^ Our deep phenotyping study investigating the metabolic and immune consequences of intermittent versus continuous NG feed delivery in healthy volunteers has highlighted a loss of the typical patterns of circulating glucose, fatty acid, triglycerides, and urea, as well as loss of the normal diurnal variation in insulin and glucagon-like peptide-1, alongside modulation of neutrophil metabolism, when feeding is delivered continuously.^29^ A study in critically ill patients found intermittent feeds resulted in a lower insulin requirement, with no additional risk of dysglycaemia.^30^

From a more pragmatic perspective there are other potential benefits to intermittent feeding as continuous NG feeding imposes restrictions on patient mobility and has to be interrupted for procedures or investigations. The frequency of these pauses in continuous feeding may explain why intermittent feeding has been shown to help reach targets for enteral calories earlier than continuous feeding.^10,12^ Additionally, a meta-analysis suggests that although intermittent feeding carries an increased risk of diarrhoea, this was balanced by a reduced incidence of constipation, with no difference in other gastro-intestinal outcomes between the groups.^31^

All of these plausible beneficial effects of intermittent feeding may improve tolerance to and recovery from critical illness. Optimising the delivery of nutrition to critically ill patients has the potential to provide several benefits: improved metabolic function with maintained insulin sensitivity; reduced catabolism and sarcopenia, which would hasten rehabilitation and improve long-term functional status; altered immune response to improve outcomes in sepsis and better entrainment of circadian rhythms with improved sleep/wake cycles, potentially resulting in reduced delirium and less risk of post-traumatic stress disorder.^32^ These benefits for patients could include a shorter, less complicated, recovery from critical illness and lower mortality. There are potential cost savings in shortened ICU and hospital stay. Crucially, implementation would not involve new drugs or technology and be straightforward and cost effective to implement as well as potentially saving staff time.

The DINE-N study aims to provide evidence to assess whether intermittent rather than continuous feed is advantageous. The study will focus on hormonal and metabolite profiles in response to intermittent versus continuous NG feeds. The primary outcome will be peak plasma insulin within 3 hours of a bolus feed compared to the equivalent time points in continuously fed patients. Insulin was chosen as the primary outcome because it is a pivotal hormone in metabolism, influencing anabolism/catabolism in the fed state, skeletal muscle autophagy, cellular energy supply, and glycaemic control.^33^ To differentiate endogenous insulin secretion from exogenous insulin administration we will simultaneously measure C-peptide.

### Aim & Objectives

The aim of this project is to establish whether intermittent feeding with overnight fasting, compared to the current standard of care in critically ill patients, produces an equivalent improvement in physiological, hormonal and metabolic responses to that seen in healthy volunteers. The research objectives will be to establish the clinical feasibility, tolerability, and efficacy of intermittent diurnal feeding in critically ill adults.

## METHODS AND ANALYSIS

### Trial Design

The study will be a prospective, parallel group, randomised, open-label trial (pre-registered at ClinicalTrials.gov: NCT06115044). The protocol for this study has been reported according to the Standard Protocol Items: Recommendations for Interventional Trials (SPIRIT) guidelines (Appendix 1).^34^ The study has received Health Research Authority (HRA) approval from Wales - Cardiff Research Ethics Committee 3 (reference 23/WA/0297). Any amendments to the current protocol will be communicated to the Research Ethics Committee via the research sponsor. The setting is the mixed ICU of Southmead Hospital, Bristol, a 996-bed teaching hospital in the southwest of England, and a major trauma centre serving an adult population of approximately 2.3 million. The ICU has 48-beds with approximately 2500 admissions annually.

### Patient and Public Involvement

A patient and public involvement (PPI) group were involved in the design of the study protocol. They informed key decisions regarding the acceptability of the intervention; blood sampling to answer the research question; and emergency waiver consent approach. One PPI representative will be included as part of the trial oversight committee to review the study conduct and outcomes.

### Population

The study will recruit a population of critically ill adult patients who are anticipated to require prolonged feeding via an NG tube (>48 hours). Participants will be recruited within 24 hours of starting enteral nutrition.

Patients must meet all the inclusion and none of the exclusion criteria as listed below.

Inclusion Criteria:

- Adults (≥18) on intensive care
- Planned for gastric enteral nutrition (anticipated duration >48 hours)

Exclusion Criteria:

- >24 hours since starting enteral nutrition
- Parenteral or jejunal nutrition
- Trophic feed only (e.g., lactate >4)
- High risk of refeeding syndrome
- Gastrointestinal surgery or pathology
- Diabetic emergencies
- Pregnancy
- Prone positioning

### Trial Intervention

The intervention is an adjustment to the normal pattern of delivery of gastric feed to deliver intermittent feeds (see Figure 1). The intermittent feeds will be administered to produce a diurnal pattern at 8:00, 13:00 and 18:00 each day. On study day 1 each feed will be 200 ml. On study day 2 each feed will be one third of total daily required volume of feed based on the normal hypocaloric targets (see Appendix 2). Each feed will be administered via volumetric pump over a period of 30 to 60 minutes. The feed type will be Nutrison Protein Plus and Nutrison Concentrated (Nutricia, UK), following standard specialist dietician-led practice.

**Figure 1.**
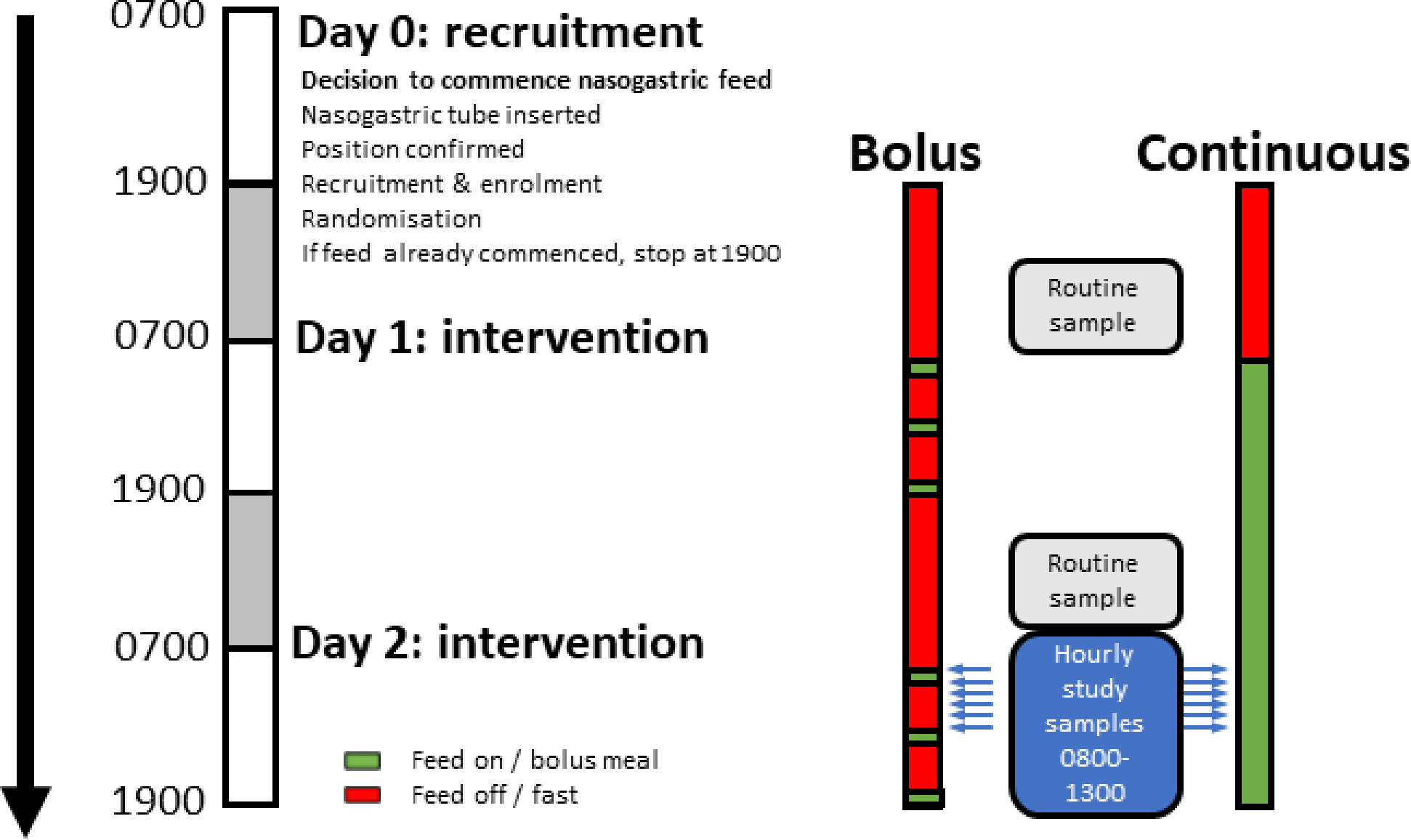
Feeding and Sampling Timeline.

The intervention period of 48 hours will start with the initiation of the overnight fast on day 0 (19:00) and will end on day 2. Patients in the intermittent feeding group will restart continuous feed at 12:00 the following day to prevent overfeeding. There will be a further 12 hours of monitoring for adverse events potentially attributable to the intervention.

### Comparator

The comparator group will have a continuous NG feeding regime that will be equivalent to the intervention group. On day of study enrolment (day 0), the continuous feed will stop at 19:00 then restart at 08:00 on study day 1 at 25 ml/hr. At 08:00 on study day 2 the rate of continuous feeding will be adjusted to meet the hypocaloric targets for ICU days 3-6 as per the local nutrition guideline (Appendix 2). At the end of the 48-hour intervention period these patients will continue on the normal continuous enteral feeding regimen.

### Outcomes

Primary outcome: The primary outcome is peak plasma insulin (and c-peptide) within 3 hours of a feed compared to the equivalent time in the continuous feed group. This will be measured for the first feed (8:00) on day 2 of the study (samples taken hourly between 8:00 and 13:00 to allow peak identification).

Secondary outcomes include:

- Endocrine and metabolic (all blood plasma)
  `o` Glucose
  `o` Ketones (beta-hydroxybutyrate)
  `o` Urea
  `o` GLP-1
  `o` Non-esterified fatty acids
  `o` Triglyceride
  `o` Glycerol
- Feasibility
  `o` % target nutrition achieved (per 24-hour period)
  `o` Protocol compliance
- Tolerability
  `o` Episodes of vomiting/24-hour period
  `o` Episodes of aspiration of feed
  `o` Delayed gastric emptying (Gastric Residual Volume >250ml x2 in a 24-hour period)
  `o` Ileus
  `o` Diarrhoea (passage of type 6 or 7 stool according to the Bristol Stool Chart or >3 stool/24 hours)
  `o` Constipation
- Efficacy
  `o` ICU and hospital length of stay
  `o` ICU and hospital mortality
  `o` Delta-SOFA (sequential organ failure assessment) between day 0 and day 2 The schedule of assessments is provided in the supplementary material (Appendix 3).

### Management of feed delivery

Reduced feed volumes will be used in all patients who are fluid-restricted or hyperkalaemic in the absence of renal replacement therapy. The study will use a standardised regime from the local nutrition guideline for assessment and management of gastric residual volumes (Appendix 2). The timing of gastric residual volume checks differs slightly from the guideline to account for the intermittent feeding pattern. During the intervention period residual volumes will be checked at 08:00, 13:00 and 18:00 with further checks overnight at 22:00, 02:00 and 06:00. Management of nausea, vomiting, diarrhoea, aspiration, ileus or constipation associated with enteral nutrition will be at the discretion of the treating intensivist.

If feed is interrupted or delayed (for example in the case of transfer for imaging or surgery) the feeding regimen will be adjusted:

- Intermittent diurnal feed: if there are two hours or more until the next scheduled feed the missed feed will be given in entirety followed by the next feed at the scheduled time. If there is less than two hours until the next scheduled feed the missed feed will be given in entirety followed by a one-hour gap and then the next scheduled feed.
- Continuous feed: the feed rate will be increased to compensate for the hours missed to achieve the prescribed 24-hour target.

### Concomitant medication

There is no restriction on concomitant medication. Enteral feed may interact with the absorption of some medications including certain antiretrovirals and antibiotics, antiepileptics and immunosuppressants. Daily medication review by a pharmacist is routine in the ICU. Alternative routes of administration and therapeutic drug monitoring will be undertaken as advised.

### Assessment of compliance

The routinely collected feeding administration record for each patient will be used to assess compliance with the intervention.

### Blood sampling

The study primary and several secondary outcomes will be measured from blood samples taken on study day 2 from 08:00 (before the start of the morning feed in the intermittent group), and at hourly intervals until and including 13:00, just before start of next feed in the intermittent group (see Figure 1). Blood samples will be taken from an indwelling catheter (10 ml per sample, arterial line or central venous line) then immediately distributed into an EDTA (ethylenediaminetetraacetic acid) and serum collection tube. All samples will be centrifuged at 3461 x g for 10 minutes for removal of the plasma or serum supernatant then frozen at -80°C pending analysis.

Analysis of samples will use standard techniques and commercially available assay kits for the relevant primary and secondary outcomes.

### Baseline Data

Baseline demographic and health related data will be collected as follows:

- Sex
- Ethnicity
- Age
- Weight
- Height
- Severity of illness (APACHE II and ICNARC physiology score)
- Organ failure assessment (Sequential Organ Failure Assessment)
- Primary reason for admission to ICU
- Time of admission
- Time of gastric tube insertion
- Time enteral feeding started prior to enrolment
- Time of enrolment
- Presence/absence of diabetes (including type if present)
- Medication (insulin, oral hypoglycaemic agents, statins)

### Assignment to Intervention

The study flow diagram is illustrated in Figure 2. Participants will be randomly allocated into two groups to receive either diurnal intermittent or continuous feed for the following 48 hours with regular monitoring. Participants will be randomised in a 1:1 ratio to intervention or control group, stratified by sex, using sealed opaque envelopes. The National Cancer Institute Clinical Trial Randomization Tool was used by a member of the ICU team independent from the study to generate the allocation sequence (https://ctrandomization.cancer.gov/). Eligibility will be confirmed, and randomisation performed, by an appropriately trained healthcare professional on the study delegation log. Participants are screened for eligibility 7 days of the week, helping to achieve an adequate rate of participant enrolment.

**Figure 2:**
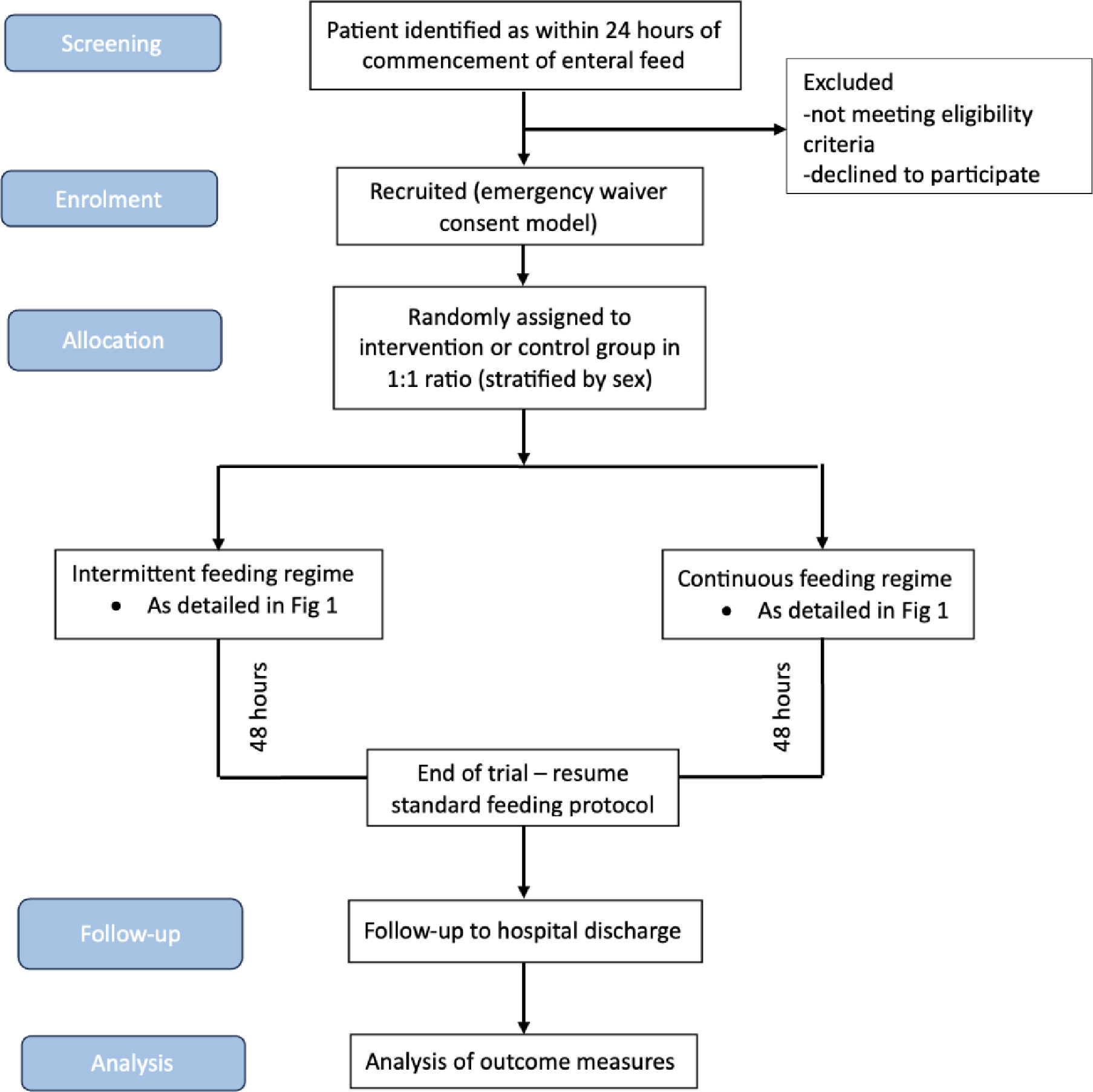
Study Flowchart.

The study is open label. Trial participants, care providers and the research team will not be masked to group allocation. The study statistician will conduct the analysis masked to group allocation.

### Consent

Informed consent will be sought from all participants. Many participants will lack capacity because of illness or the required interventions. Owing to the time-critical nature of enteral feeding, and as prolonged feeding prior to intervention may bias the study, it is not practicable to wait until capacity returns. Seeking Personal Consultee opinion in an emergency may cause additional distress for relatives. As such, an emergency waiver of consent model will be used, with informed consent sought once patients regain capacity (Appendix 4). Participants (or their Personal or Professional Consultee) are free to withdraw at any time from the study without giving reasons and without prejudicing any further treatment. This approach was deemed appropriate in a PPI consultation. See Appendix 5 for the participant information sheet approved by the Research Ethics Committee.

The biological samples will be used only for the pre-defined schedule of assessments in this study, and then discarded. The data generated from samples can be used for ancillary studies.

### Participant study exit criteria

The trial intervention will be terminated if any of the following events occur (within 48 hours of study entry):

- Death
- Withdrawal of life sustaining treatment
- Early discharge from the ICU
- Decision by attending clinician that the delivery of feed should be stopped or adjusted on safety grounds
- Development of any of the conditions listed in the exclusion criteria (i.e., need for prone positioning or trophic feeding)

### Post-trial care

After the trial intervention period, participants will be managed and monitored following the local ICU unit guidelines in general and specifically for nutrition (Appendix 2).

### Participant Withdrawal

Participants, or their personal or professional consultees, may withdraw from the study at any time. No reason needs to be given and usual medical care will not be affected. Only data essential for study monitoring and oversight will be retained.

### End of trial

The trial will end upon completion of the follow-up period for the final participant or if required by the Sponsor, Research Ethics Committee, or the Trial Oversight Committee.

### Data collection, management, and analysis

Data will be collected on a paper case report form (CRF) according to the schedule assessments outlined in the Appendix. All missing data must be explained on the CRF. Paper copies of the CRF will be kept in a secure location (locked cabinet). An online Redcap database will be used to store and assimilate clinical and assay data. All documents will be stored securely and only accessible by trial staff and authorised personnel. Data will be collected and retained in accordance with the relevant data protection legislation. Access to the data will be granted to authorised representatives from the Sponsor, host institution and the regulatory authorities to permit trial-related monitoring, audits and inspections.

Retention in the study is promoted by 7-day research nurse support and education of clinical teams. Follow-up for ICU and hospital outcomes uses routinely collected national clinical audit data. No distinction is made in data collection between participants who deviate or discontinue from intervention protocols.

### Access to the final trial dataset

The co-applicants, collaborators and Sponsor will have access to the final trial dataset. Applications for access to the final trial dataset will be considered by the Chief Investigator and Sponsor after publication of trial results.

### Statistical Analysis

In a study of healthy participants, the intermittently fed group had a mean (SD) peak plasma insulin concentration at 2 hours after the bolus of 373±204 pmol/l compared to the continuous feed group 58±41 pmol/l.^29^ This effect size of 2.14 would require 6 subjects per group to have a 90% power (p=0.05). We expect a smaller effect size in the critically ill population as the volume of bolus feeds are less than in the healthy participant studies and they are likely to have less physiological capacity to mount a response. In addition, a smaller response in critically ill patients would be clinically relevant and there is likely to be more variability in the data given the heterogeneity in the patient sample and the potential for violation of the assumption of equal variances. We have therefore adjusted the sample size calculation to be able to detect a smaller effect of 1.26 whereby the study would have 80% power with 11 patients per group, and 90% power with 15 patients per group. The sample size has been adjusted to 30 to detect this more conservative effect size estimate while still retaining at least 80% power in the case of a 25% drop out rate.

All data will be analysed using an intention-to-treat analysis set. Analysis will be undertaken by the study statistician, masked to group allocation. A full review of the data (data veracity, verification and validity) will be undertaken prior to inferential analysis. If the amount of missing data on an outcome is between 20-40% we will carry out a sensitivity analysis under ‘Missing Not At Random’ scenarios. If the amount of missing data on an outcome is >= 40%, these data will be reported descriptively.

Two-sided statistical tests will be used throughout and a p-value < 0.05 will be taken as statistically significant. The primary outcome is peak plasma insulin within 3 hours of the 2^nd^ day morning bolus feed compared to the equivalent time point in continuously fed patients. A robust comparison of means on the primary outcome will be undertaken using the most appropriate form of the two-sample t-test (independent samples, Welch test, or bootstrap equivalent if there is a severe violation of underpinning assumptions). Effect size will be reported using 95% confidence intervals. The same analyses at the 3 hours of bolus feed compared to the equivalent time point in continuously fed patients will be undertaken for hormonal and metabolic indicators (C-peptide, fatty acid, glycerol, triglyceride, GLP-1, glucose, ketones, urea). Area under the curve of hormonal and metabolic measures tested hourly on study day 2 will be compared between groups using the same methodology.

Haematology, biochemistry, and acid-base balance measures derived from routine daily blood sampling will be compared between randomised arms for each day using the above methodology, assessed for changes over time using the paired samples t-test, and with change scores assessed between randomised arms. There are no planned interim analyses.

Kaplan-Meier analyses coupled with the log-rank test will be used to compare the length-of-stay outcomes between randomised arms. The percentage achieving target nutrition (per 24-hour period) and protocol compliance will be reported by randomised arm with 95% CI for between group differences.

### Monitoring

The study will be monitored in accordance with local hospital guidelines according to a plan agreed by the sponsor. All trial-related documents will be made available on request for monitoring and audit by the Research Sponsor, Research Ethics Committee and for inspection by the Medicines and Healthcare products Regulatory Authority or other licensed bodies. A formal data monitoring committee was not considered to be required given the limited scope of the study. The Trial Management Group consisting of the study investigators and Sponsor’s representative meets quarterly to review study progress. The Oversight Committee of at least two independent clinicians and one PPI representative will review the reports and recommendations of the trial management group. Any suspected adverse events will be reported to the study sponsor.

### Dissemination

The trial will be reported in accordance with the Consolidated Standards of Reporting Trials (CONSORT) guidelines (www.consort-statement.org). The main report will be written by the Trial Management Group with authorship determined according to the internationally agreed criteria for authorship (www.icmje.org).

The study will be presented at scientific and clinical meetings and uploaded to a pre-print server prior to publication in an open access peer-reviewed journal. Participants will be asked if they wish to have a lay summary of the findings.

## DISCUSSION

Despite the widely recognised importance of adequate nutrition for a large proportion of the critically ill population, there is a shortage of evidence comparing outcomes between different feeding strategies and specifically between continuous and intermittent regimes. The existing trials are small (typically less than 100 participants in each arm) and implement a variety of intermittent feeding regimes, which have been difficult to synthesise in meta-analyses. Many studies fail to delineate adequate fasting periods, particularly overnight, which limits the ability to contrast with continuous feeding regimes (as gradual stomach emptying likely smooths out the delivery of nutrients to the gut). Recent reviews highlight the paucity of studies looking at metabolic and hormonal outcomes from intermittent diurnal feeding.^31,35^ We detected differences in metabolic and immune profiles in healthy volunteers, but it is not yet known whether these findings translate to the ICU population, whose physiology may be altered by critical illness. This study will start to address this knowledge gap in the literature by focusing on the metabolic and hormonal effects of continuous vs. intermittent NG feeding.

The potential for intermittent feeding to increase the risk of adverse gastro-intestinal outcomes continues to be a source of controversy. There is suggestion of a trend towards increased incidence of constipation in continuous feeding groups, and increased incidence of diarrhoea in intermittent feeding groups. This study will contribute towards clarifying the side-effect profile for continuous and intermittent feeding regimes. Even if there are differences this may indicate the need to individualise the choice of different feeding patterns depending on patient factors.^36^

A strength of the study includes the involvement of dieticians, specialists in nutrition and metabolism, and patient and public involvement in the research design and delivery. The eligibility criteria are broad, which should help to generate a representative sample of the ICU patient population. The intermittent feeding intervention in this study has been carefully considered, and involves sufficient breaks between feeds, with an overnight fast, to allow us to capture feeding/fasting cycles in metabolites. Hourly blood sampling around the time of a meal represents an innovative approach that allows detailed profiling of metabolic hormones and will increase our ability to detect a difference between continuous and intermittent feeding regimes.

We anticipate several limitations to this study. An open-label design has been chosen, which may bias the interpretation of the results. Only a single centre will be studied, limiting the generalisability and external validity of the study. Despite intensive blood sampling, there is a chance that, by only sampling on day 2 of the trial period, we may fail to detect differences in metabolism that are present at different time points. In addition, it is understandably not feasible to undertake any more invasive tissue/mechanistic measurements in this exploratory study.

This study will unveil detailed information on the effects of intermittent versus continuous feeding on metabolic hormone profiles. We aim to use the study findings to inform the design of a multicentre RCT powered to demonstrate the efficacy of different nasogastric feeding regimes on length of stay, mortality, and important long-term patient-centred outcomes such as physical function. This preliminary study may highlight benefits of intermittent feeding regime from a practical standpoint, for example in terms of patient mobility and acceptability to patients and staff, so consideration will be given to a definitive mixed methods study design to investigate these factors. Any benefits of the intermittent regime may also be translated to resource-poor settings where the limited availability of pumps may prevent continuous administration.

### Conclusion

Here we present the protocol for a proof-of-concept study to investigate whether there are different metabolic outcomes between continuous and intermittent feeding regimes in critically ill patients. This study will offer novel insight into important circadian, hormonal, and metabolic responses to different feeding regimes.

### Trial Status

Recruiting

### Protocol Version

1.3 28^th^ September 2023 (Appendix 6)

## Data Availability

All data produced in the present work are contained in the manuscript

## Author’s Contributions

JB, HS, JG, TP, MA, MT, DM conceived and designed the study; CB, AR, BB, KH, MT, DM oversaw delivery and data collection; PW will lead statistical analysis. All authors have reviewed and approved the final manuscript.

## Funding and Sponsor

This study is funded by the Southmead Hospital Charity Research Fund. TEP and MA are supported by MRC funding for studies of metabolism related to ICU care (MR/W029138/1). North Bristol NHS Trust is the sponsor of the trial and contact details are given below. Neither funder nor sponsor have a role in study design, delivery, analysis, interpretation or decision to submit for publication.

Sponsor Contact Information:

North Bristol NHS Trust

Research & Innovation

Southmead Hospital, Bristol, BS10 5NB

## Supplementary Material

Appendix 1. SPIRIT Checklist

Appendix 2. Local ICU Nutrition Guideline

Appendix 3. Schedule of Assessments

Appendix 4. Consenting Process

Appendix 5. Participant Information Sheet

Appendix 6. Protocol

