## Appendix 2 for "Does Intermittent Nutrition Enterally Normalise hormonal and metabolic responses to feeding in critically ill adults? A protocol for the DINE-Normal proof-of-concept randomised parallel group study"

### Intensive Care Unit Nutrition Guideline

This guideline is a quick reference guide to feeding patients on ICU at North Bristol NHS Trust.

| Version 1.1 | Valid from 01/03/2022 | Review due 01/03/2024 | Authors: Stephen Taylor, Rowan Clemente |
|---|---|---|---|

#### Introduction

This guideline applies to all patients admitted to Southmead ICU. Follow the enteral feeding process diagram on page 2 for all patients.

#### Aims and rationale

##### Days 1-2

Support physiological adaptation to critical illness by:

- Correcting micronutrient deficit to optimise metabolic and anti-oxidant systems.
- Provide minimal EN macronutrients to maintain GI function and immunity.

##### Days 3-6

Hypocaloric, high nitrogen feeding:

- Energy expenditure: Provide <60% if obese, <80% if other to avoid substrate intolerance
- Nitrogen: 0.2-0.32g/kg/day to optimise wound healing and acute-phase protein response.

##### Day 7 onwards

Meet full requirement.

- Energy: Dietitian judges when to meet energy expenditure as substrate tolerance permits.
- Nitrogen and bolus feed/ food: Time to optimise activity-induced anabolism.

#### Enteral feeding decision tree

Following admission to ICU, the flow chart on the next page should be followed for all patients (excluding PACE admissions).


*This document can only be guaranteed to be the current adopted version if opened directly from the NBT intranet.*
Exceptional healthcare, personally delivered

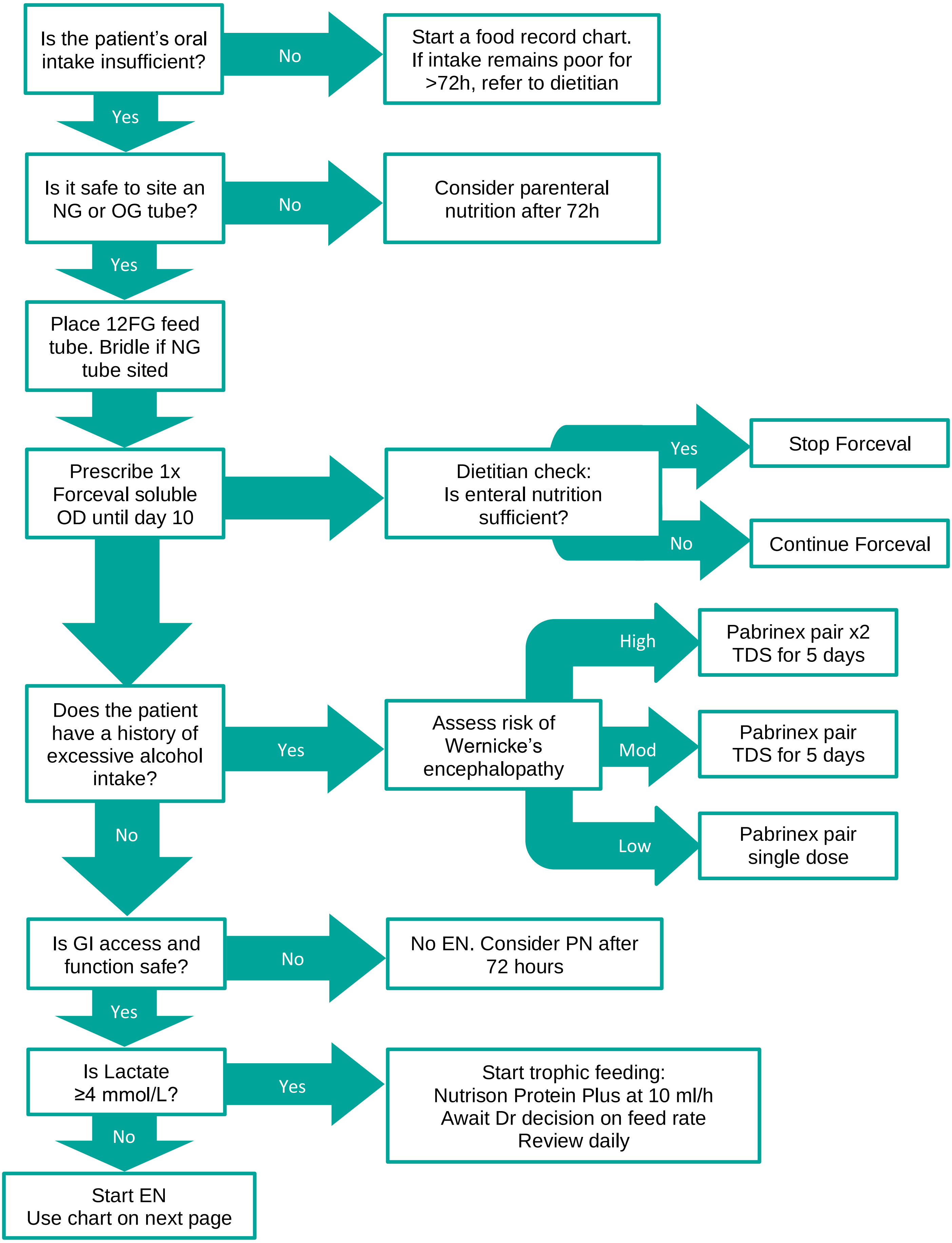
Is the patient's oral intake insufficient?
No
Start a food record chart. If intake remains poor for >72h, refer to dietitian
Yes
Is it safe to site an NG or OG tube?
No
Consider parenteral nutrition after 72h
Yes
Place 12FG feed tube. Bridle if NG tube sited
Prescribe 1x Forceval soluble OD until day 10
Dietitian check: Is enteral nutrition sufficient?
Yes
Stop Forceval
No
Continue Forceval
Does the patient have a history of excessive alcohol intake?
Yes
Assess risk of Wernicke's encephalopathy
High
Pabrinex pair x2 TDS for 5 days
Mod
Pabrinex pair TDS for 5 days
Low
Pabrinex pair single dose
No
Is GI access and function safe?
No
No EN. Consider PN after 72 hours
Yes
Is Lactate ≥4 mmol/L?
Yes
Start trophic feeding: Nutrison Protein Plus at 10 ml/h Await Dr decision on feed rate Review daily
No
Start EN Use chart on next page

#### Nutrition action plan

The following decision tool is designed to summarise responsibilities of nursing and medical staff following admission along with providing an aide-memoire for enteral feeding rates and phosphate replacement.

| | | Screen | Action | | | | Daily WR review |
|---|---|---|---|---|---|---|---|
| ADMISSION | Nurse | **Insufficient food<br>NGT in situ** | Place 12F NGT if necessary and possible<br>Bridle, ensure clip 0.5cm from septum | | | | Confirm tube position |
| ADMISSION | Doctor | **All patients** | Forceval soluble | 1 tablet OD NG until day 10 | | | Dietitian may cancel |
| ADMISSION | Doctor | **Wernicke's risk** | | **High** | **Possible** | **Low** | Symptom > dose review |
| | | | Pabrinex 1 pair | 2 TDS | 1 TDS | 1 OD | |
| | | | Duration | 5 days | 5 days | One off | |
| ADMISSION | Doctor | **Burn, CRRT** | 1 pair Pabrinex IV OD & 10mL Additrace IV OD | | | | Dietitian will review |
| ADMISSION | Doctor | **No GI access / poor function** | No enteral nutrition | | | | **Consider** TPN after 72h |
| ADMISSION | Doctor | **Lactate > 4.0** | 10ml/h Nutrison Protein Plus until WR decision. | | | | Feed rate decision |

| | | | | | | | |
|---|---|---|---|---|---|---|---|
| DAILY | Nurse | **Start NG feed (ml/hr)** | **Nutrison** | **Most patients**<br>**Protein Plus** | **Fluid restricted or K+ >5.0 & no CRRT**<br>**Concentrated** | | **Check gastric residual volume 4 hourly** |
| | | | Day 1 & 2 | 30 | 20 | | < 250mL bile/feed: |
| | | | Then: full feed | Dietitian regime or use actual weight (kg) | | | ➔ Return + full feed rate |
| | | | 40kg | 40 | 27 | | ⬇ |
| | | | 50kg | 45 | 30 | | ≥ 250mL or blood / faecal / vomit |
| | | | 60kg | 50 | 32 | | ➔ Discard + full feed rate |
| | | | 70kg + | 55 | 35 | | ⬇ |
| DAILY | Doctor | **Phosphate** | **IV polyfusor** | **ml** | **ml/hr** | **Hours** | If 2nd > 250mL or vomit<br>➔ Metoclopramide 10mg IV TDS |
| | | < 0.5 * | | 400 | 33 | 12 | ⬇ |
| | | < 0.65 * | | 300 | 25 | 12 | 24h: unresolved or ≤ full rate EN? |
| | | | * If <72h of feed > ⬇ feed to 30ml/h until phosphate > 1.0 | | | | ⬇ |
| | | <0.8 | | 200 | 17 | 12 | Request NJ via dieticians |
| | | 0.8-1.0 or if previous day <0.8 | **Phosphate sandoz** | 1 tablet TDS | | | If > 24h delay for NJ:<br>Erythromycin 250mg IV QDS |
| | | **CRRT** | **Adjust daily supplement to maintain PO4 at 1.0-1.4** | | | | Version 2.0, December 19 |
| | | **New infusion** | **Recheck phosphate level before commencing** | | | | |

#### Management of gastric aspirates

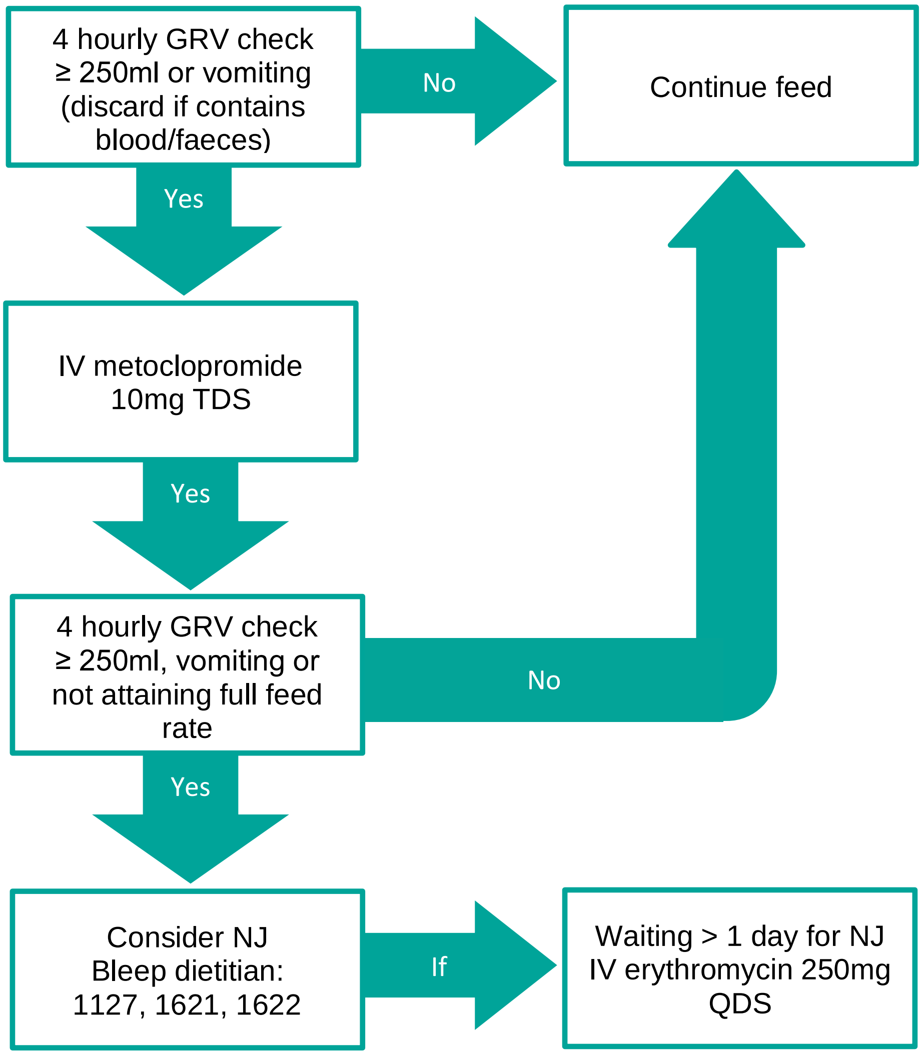


#### Total Parenteral Nutrition (TPN)

##### In hours

If the multidisciplinary team have made the clinical decision that TPN is required, bleep your Pod dietitian.

##### Out of hours

If the multidisciplinary team have made the clinical decision that TPN is required over the weekend:

- Complete the TPN calculator (intranet) to determine safest infusion rate

| Responsibility | Name | Division / Specialty | Job Title |
|---|---|---|---|
| **Authorised by** | **ICU SDM** | Intensive Care Unit | - |
| **Author** | Stephen Taylor | Intensive Care Unit | Dietitian |
| **Author** | Rowan Clemente | Intensive Care Unit | Dietitian |
