## Appendix 3 for "Does Intermittent Nutrition Enterally Normalise hormonal and metabolic responses to feeding in critically ill adults? A protocol for the DINE-Normal proof-of-concept randomised parallel group study"

**APPENDIX - Schedule of Assessments**

| **Assessment** | **Measurement** | **Timepoint(s) of evaluation** |
| --- | --- | --- |
| Hormonal and metabolic | Insulin, C-peptide, fatty acid, glycerol, triglyceride, GLP-1, glucose, ketones urea | Tested hourly on Study Day 2 as per research bloods (Figure 2) |
| Acid-base balance | Arterial blood gas (pH, bicarbonate, lactate, base excess) | With routine daily blood sampling |
| Haematology | Haematocrit, haemoglobin, platelet count, red cell distribution width, red blood cell count, white blood cell count, neutrophils, lymphocytes.  Activated partial thromboplastin time, prothrombin time, International Normalised Ratio (INR)  C-reactive protein | With routine daily blood sampling |
| Biochemistry | Albumin, alkaline phosphatase, alanine aminotransferase, calcium, creatinine, magnesium, phosphate, potassium, sodium, total bilirubin, urea, eGFR | With routine daily blood sampling |
| Clinical | SOFA  Mortality and length of stay | Baseline, day 2  ICU and hospital discharge |
| Nutrition | Vomiting  Diarrhoea  Delayed gastric emptying  Ileus  Aspiration  Total enteral nutritional intake | During study intervention period |
