## Appendix 4 for "Does Intermittent Nutrition Enterally Normalise hormonal and metabolic responses to feeding in critically ill adults? A protocol for the DINE-Normal proof-of-concept randomised parallel group study"

**APPENDIX - DINE-Normal Consenting**

**1. Emergency Waiver**

- Once eligibility has been confirmed, a verbal explanation will be given to the Personal Consultee if available and appropriate.
- If no Personal Consultee is available/ a discussion inappropriate due to distress, the participant may be enrolled under emergency waiver.
- Personal Consultees will be given/ offered an information sheet if available at emergency waiver stage.
- An emergency waiver form is completed by the research team, one copy will be entered into the medical notes and the original remains in the ISF
- Should the consultee decline following discussion, the participant is entered onto the screening log and not enrolled.

**2. Personal Consultee**

- Trial procedures continue while a Personal Consultee is sought.
- Once an appropriate consultee is identified, a full explanation is given by the research team with Personal Consultee Information Sheet.
- Adequate time should be given for the consultee to consider participation – this should be no less than 1 hour. Although a guideline of 24 hours is advised, there is no upper limit to time for consideration.
- The Personal Consultee Opinion form is signed by the representative and a member of the research team with this delegated task. A copy is given to the representative; a copy is place in the medical notes, and the original to the ISF.
- If a process of Personal Consultee has commenced in ICU, this should be continued into the ward setting. However, if the patient is transferred to a ward or another hospital prior to commencing Personal Consultee, the process may be deferred to Professional Consultee.

**3. Professional Legal Representative**

- Trial procedures continue while a Professional/ Personal Consultee are sought.
- If a Personal Consultee is not identified within 72 hours, a consultant unconnected with the study or an Independent Mental Capacity Advocate may act as a Professional Consultee.
- A Professional Consultee Opinion form must be signed if a Personal Consultee has not been identified, or if it remains inappropriate to approach.
- A Professional Consultee Opinion form must be signed prior to patient transfer if this occurs.
- The Professional Consultee Opinion form is signed by the representative and a member of the research team with this delegated task. A copy is given to the representative; a copy is place in the medical notes, and the original to the ISF.
- If a Personal Consultee is subsequently identified, Personal Consultee Opinion should be sought and supersedes the opinion of the Professional Consultee opinion.

**4. Retrospective Patient Consent**

- The participant is observed for regaining capacity to consent for the duration of the hospital stay.
- If the patient has regained capacity for full understanding, consent is sought by the research team with the delegated task and supersedes the opinion of both Personal and Professional Consultee Opinions
- Adequate time should be given for the patient to consider continued participation – this should be no less than 1 hour. Although a guideline of 24 hours is advised, there is no upper limit to time for consideration.
- Participants, or their personal or professional consultees, may withdraw from the study at any time. No reason need be given, and usual medical care will not be affected. Only data essential for study monitoring and oversight will be retained.
