## Appendix 5 for "Does Intermittent Nutrition Enterally Normalise hormonal and metabolic responses to feeding in critically ill adults? A protocol for the DINE-Normal proof-of-concept randomised parallel group study"

**Does intermittent enteral feeding engage post-prandial hormonal, digestive and metabolic responses in critically ill patients? A proof-of-concept study.**

Chief Investigator: Matt Thomas

**Section 1 - Participant Information Sheet**

This information sheet gives you details of a study we are running in the Intensive Care Unit in Southmead Hospital. Before you decide whether you wish to continue to be involved in this study, it is important for you to understand why this research is being done and what it would involve for you.

We understand that you have been through an extremely difficult time and hope that giving you more information will not add to any distress that have experienced. Please take time reading this information and make note of questions as you read. One of our team will be happy go through this information with you and answer any questions you may have. You may wish to talk to your friends and family about this study before you decide if you would like to continue to take part.

The purpose of the study is to determine whether there may be a benefit to changing the timing of artificial feeding in critically ill patients. Currently, approximately half of patients in the Intensive Care Unit need artificial feeding which is normally delivered continuously throughout the day and night. An alternative feeding pattern, where the feed is given as three separate “meals” during the day, may be better for critically ill adults.

**Patient information and health and care research**

The Health Research Authority and government departments in Northern Ireland, Scotland and Wales set standards for NHS organisations to make sure they protect privacy and comply with the law when they are involved in research. The Research Ethics Committee has reviewed this research study to make sure that the use of data is in the public interest and meets ethical standards.

Health and care research may be exploring prevention, diagnosis or treatment of disease, which includes health and social factors. Research may be sponsored by companies developing new medicines or medical devices, NHS organisations, universities or medical

research charities. The research sponsor decides what information will be collected for the study and how it will be used.

Health and care research should serve the public interest, which means that research sponsors must demonstrate that their research serves the interests of society as a whole. They do this by following the [UK Policy Framework for Health and Social Care Research](https://www.hra.nhs.uk/planning-and-improving-research/policies-standards-legislation/uk-policy-framework-health-social-care-research/). They also must have a legal basis for any use of personally identifiable information.

**Why are we doing this research?**

We are doing this study because members of our team have shown giving artificial feed continuously to healthy people produces a very abnormal response in the body. As giving artificial feed continuously is usual when people are very sick, we would be worried if the same was true for patients in the Intensive Care Unit. Giving artificial feed in a more natural pattern as separate meals with a fast overnight (“intermittent”) could have benefits for people who are sick enough to end up in an Intensive Care Unit. This study is the first step to finding out if this is true.

**What is already known?**

In both humans and animals evidence suggests that intermittent feeding aligned with natural body rhythms improves circulation to the gut and promotes pulses in crucial digestive hormones. Intermittent feeding has also been shown to have benefits to the immune system and digestion of protein, and these effects could translate to better recovery from critical illness.

**Why was I chosen?**

You were chosen because you were very unwell on the Intensive Care Unit and needed to have artificial feed through a tube into your stomach.

**What has happened to me so far?**

After you were admitted to the Intensive Care Unit a feeding tube was inserted into your stomach via your nose (this is part of routine care for patients). Artificial feed was given through this feeding tube in one of two patterns. In half the patients in the study the feed is given continuously, as standard. In the other half it is given as three “meals” with a fast overnight. Apart from the modified feeding schedule, all other care during the 48-hour period is usual standard of care. Blood samples have been taken to investigate the effect on insulin and other hormones.

Most patients who are very sick in the Intensive Care Unit are anaemic (have a low haemoglobin blood count). A small minority of patients need a blood transfusion. The amount of blood taken for this study (approximately 4tablespoons) is not enough to make this more likely.

**Why am I being asked after the study has happened?**

As you were very ill in the Intensive Care Unit it was not possible to ask you at the time for your agreement. Asking permission from a relative or a friend would have caused a delay and we know starting feed early is vital. However, we will have already discussed this research study with a relative, friend or hospital consultant when it was possible. We have now come to talk to you as soon as possible, and when you feel able to read this information; this method of consent is commonly used in emergency studies.

**What happens to me during the rest of the study?**

The study period is only 48 hours. After that is over all care is as standard for patients in the Intensive Care Unit and on the hospital ward.

**What will happen to the samples I give?**

The blood is processed and stored at North Bristol NHS Trust before transfer to the University of Bath for analysis for insulin and other hormones. The information attached to the samples will not identify you and will not be combined with other information in a way that could identify you. The samples will only be used for the purpose of this research and cannot be used to contact you or to affect your care. Once the study ends the samples will be destroyed within 5 years, in accordance with the Human Tissue Act 2004. If you decide to withdraw from the study, you can ask for samples that have not been analysed to be destroyed. If the samples have been analysed the results will not be used for the study.

**Do I have to continue to take part?**

It is up to you if you wish to continue to take part. You are free to leave the study at any time without giving a reason, and this does not affect your medical care now or in the future. If you choose to continue to take part, one of the research team will discuss the consent form with you and address any questions you may have.

**What are the possible benefits of taking part?**

We cannot promise that you will get any benefit from taking part in this study. The information we obtained from your participation in the study may inform us about future

treatments for tens of thousands of patients each year who have artificial feeding in an Intensive Care Unit.

**What are the possible disadvantages and risks of taking part?**

We know that stomach and bowel upsets, including vomiting, diarrhoea and constipation, are very common in patients in the Intensive Care Unit. We do not know if the intermittent feeding pattern will make this any worse.

For some patients who are vomiting there is a risk of contaminating the lungs (“aspiration”) which can make people even sicker. We do not know if the intermittent feeding pattern will make this risk any higher.

Patients who are very unwell may have low or more often high blood sugar. We do not know if the intermittent feeding pattern will make this more likely.

**What if something goes wrong?**

If it appears that you are suffering any serious ill-effect from your participation in the study, you will be withdrawn from the study immediately.

In the unlikely event that you are harmed as a result of your participation in the study, you will be compensated, without having to prove negligence, provided that on balance of probabilities, an injury was caused as a direct result of the procedures you received during the course of the study. These special compensation arrangements apply where an injury is caused that would not have occurred if you were not in the trial. If you are harmed due to someone’s negligence, then you may have grounds for legal action, but you may have to meet the legal costs. The normal NHS complaints mechanisms will still be available.

**What will happen if I don't want to carry on with the study?**

If you do not want to carry on with the study, we will withdraw you and you will take no further part. You are under no obligation to continue in the study. If you decide to withdraw, we will not use any data from you for the results of the study.

**Section 2 – General Data Protection Regulation**

**What is patient data?**

When you go to your GP or hospital, the doctors and others looking after you will record information about your health. This will include your health problems, and the tests and treatment you have had. They might want to know about family history, if you smoke or what work you do. All this information that is recorded about you is called patient data or patient information.

When information about your health care joins together with information that can show who you are (like your name or NHS number) it is called identifiable patient information. It’s important to all of us that this identifiable patient information is kept confidential to the patient and the people who need to know relevant bits of that information to look after the patient. There are special rules to keep confidential patient information safe and secure.

**What sort of patient data does health and care research use?**

There are lots of different types of health and care research. The research team will be looking at some of the patient’s health records. This sort of research may use some data from their GP, hospital or central NHS records. Some research will combine these records with information from other places, like schools or social care. The information that the researcher collects from the health records is research data.

**Why does health and care research use information from patients?**

In clinical trials, the researchers are collecting data that will tell them whether one treatment is better or worse than other. The information they collect will show how safe a treatment is, or whether it is making a difference to your health. Different people can respond differently to a treatment. By collecting information from lots of people, researchers can use statistics to work out what effect a treatment is having.

**How does research use patient data?**

In this research study we will use information from your clinical records. We will only use information that we need for the research study. We will let very few people know your name or contact details, and only if they really need it for the study. You have been assigned a unique code by which any samples and results will be recorded. Your name or any other personal information will not be directly associated with any samples or results. An electronic document linking your details to your study code will be stored in an encrypted file on a password protected computer system accessible only to the study team.

**Where will my data go?**

In this study, the research team will collect your data from and store your data within the NBT and NHS secure network. Everyone involved in this study will keep your data safe and secure. We will also follow all privacy and GDPR rules. The identifiers for each participants’ data will be kept until the results are available. The results of this study will be discussed among the study team at the end of the study, presented at scientific meetings and used to produce academic publications. You cannot be identified in any of the results or publications. Anonymised data will be kept in a secure data archive for 5 years after the study has ended as per standard protocol.

**What if I don't want my patient data used for research?**

You will have a choice about taking part in a clinical trial testing a treatment. If you choose not to take part, that is fine.

In most cases you will also have a choice about your patient data being used for other types of research. There are two cases where this might not happen:

- When the research is using anonymous information. Because it’s anonymous, the research team don’t know whose data it is and can’t ask you.
- When it would not be possible for the research team to ask everyone. This would usually be because of the number of people who would have to be contacted. Sometimes it will be because the research could be biased if some people chose not to agree. In this case a special NHS group will check that the reasons are valid. You can opt-out of your data being used for this sort of research. You can ask your GP about opting-out, or you can find out more about how we use your information:
- at [www.hra/nhs.uk/information-about-patients/](http://www.hra/nhs.uk/information-about-patients/)
- our leaflet available from [www.nbt.nhs.uk/PatientResearchdata](http://www.nbt.nhs.uk/PatientResearchdata)
- by asking one of the research team
- by contacting Helen Williamson (Head of Information Governance) at or by ringing 01174144767

**Who is organising and funding this study?**

This research is being sponsored by North Bristol NHS Trust. The study is funded by the Southmead Hospital Research Fund Charity.

**Who has reviewed this study?**

All research in the NHS is assessed by an independent group of people, called a Research Ethics Committee (REC), to protect your interests. This study has been reviewed and approved by ***********.

**Further information and contact details**

Further information can be gained from:

Dr Matt Thomas (Chief Investigator)

North Bristol NHS Trust Research and Innovation (Sponsor)

**Thank you for taking the time to read this. Please ask if there are any questions.**
