## Appendix 6 for "Does Intermittent Nutrition Enterally Normalise hormonal and metabolic responses to feeding in critically ill adults? A protocol for the DINE-Normal proof-of-concept randomised parallel group study"

**FULL/LONG TITLE OF THE TRIAL**

**D**oes **I**ntermittent **N**utrition **E**nterally **Normal**ise hormonal and metabolic responses to feeding in critically ill adults? The DINE-Normal proof-of-concept study.

**SHORT TRIAL TITLE / ACRONYM**

The DINE-Normal proof-of-concept study.

**PROTOCOL VERSION NUMBER AND DATE**

Version 1. Final 28 September 2023

**This protocol has regard for the HRA guidance and order of content**

**RESEARCH REFERENCE NUMBERS**

IRAS Project ID:	328469

Trial registration:	ClinicalTrials.gov

Sponsor Number:	R&D 5454

**SIGNATURE PAGE**

The undersigned confirm that the following protocol has been agreed and accepted and that the Chief Investigator agrees to conduct the trial in compliance with the approved protocol and will adhere to the principles outlined in the GCP guidelines, the UK Policy Framework for Health and Social Care Research, the Sponsor’s (and any other relevant) SOPs, and other regulatory requirements as amended.

I agree to ensure that the confidential information contained in this document will not be used for any other purpose other than the evaluation or conduct of the clinical investigation without the prior written consent of the Sponsor

I also confirm that I will make the findings of the trial publicly available through publication or other dissemination tools without any unnecessary delay and that an honest accurate and transparent account of the trial will be given; and that any discrepancies and serious breaches of GCP from the trial as planned in this protocol will be explained.

| **For and on behalf of the Trial Sponsor:** | | |
| --- | --- | --- |
| Signature:  ...................................................................................................... |  | Date: ....../....../...... |
| Name (please print):  ...................................................................................................... |  |  |
| Position: ...................................................................................................... |  |  |
| **Chief Investigator:** | | |
| Signature: ...................................................................................................... |  | Date: ....../....../...... |
| Name: (please print):  ...................................................................................................... |  |  |

### KEY TRIAL CONTACTS

| Chief Investigator | Dr Matt Thomas  Intensive Care Unit  Southmead Hospital  Bristol BS10 5NB   0117 950 5050 |
| --- | --- |
| Principal Investigator | Dr Clodagh Beattie  Intensive Care Unit  Southmead Hospital  Bristol BS10 5NB   0117 950 5050 |
| Trial Administrator | Clinical Research Centre  Southmead Hospital  Bristol BS10 5NB  0117 414 8136 |
| Sponsor | North Bristol NHS Trust  Research & Innovation  Southmead Hospital  Bristol BS10 5NB   0117 414 9333 |
| Funder(s) | Southmead Hospital Charity Research Fund  Southmead Hospital Intensive Care Unit |
| Key Protocol Contributors | Dr Matt Thomas  Professor Tony Pickering  Dr Mike Ambler  Ms Danielle Milne  Dr Clodagh Beattie  Professor James Betts  Mr Harry Smith  Professor Javier Gonzalez  Professor Paul White  Dr Aravind Ramesh  Ms Borislava Borislavova |
| Statistician | Dr Paul White  Department of Engineering Design and Mathematics  University of the West of England  Bristol BS16 1QY .ac.uk  0117 328 3777 |

i. **CONTENTS**

12	[MONITORING, AUDIT & INSPECTION……………………………………………………….34](#_Toc490143095)

**ii. LIST OF ABBREVIATIONS**

ACBS	Advisory Committee on Borderline Substances

AE	Adverse Event

ANOVA	Analysis of variance

AR	Adverse Reaction

APACHE	Acute Physiology and Chronic Health Evaluation

CG	Clinical Guideline

CI	Chief Investigator

CRF	Case Report Form

CTA	Clinical Trial Authorisation

CTIMP	Clinical Trial of Investigational Medicinal Product

DSUR	Development Safety Update Report

EDTA	Ethylenediaminetetraacetic acid

eGFR	Estimated glomerular filtration rate

EN	Enteral nutrition

GCP	Good Clinical Practice

GLP-1	Glucagon-like peptide 1

GRV	Gastric residual volume

HRA	Health Research Authority

ICCA	Intellispace Critical Care and Anaesthesia

ICF	Informed Consent Form

ICH-GCP	International Committee on Harmonisation – Good Clinical Practice

ICNARC	Intensive Care National Audit and Research Centre

ICU	Intensive Care Unit

INR	International Normalized Ratio

IMP	Investigational Medicinal Product

ISF	Investigator Site File

ISRCTN	International Standard Randomised Controlled Trials	Number

MA	Marketing Authorisation

MHRA	Medicines and Healthcare products Regulatory Agency

NBT	North Bristol NHS Trust

NG	Nasogastric

NHS	National Health Service

NHS R&D	National Health Service Research & Development

NICE	National Institute for Health and Care Excellence

OG	Orogastric

PI	Principal Investigator

PIS	Participant Information Sheet

REC	Research Ethics Committee

SAE	Serious Adverse Event

SAR	Serious Adverse Reaction

SDV	Source Data Verification

SOFA	Sequential Organ Failure Assessment

SOP	Standard Operating Procedure

SmPC	Summary of Product Characteristics

SSI	Site Specific Information

SUSAR	Suspected Unexpected Serious Adverse Reaction

TMF	Trial Master File

TMG	Trial Management Group

TOC	Trial Oversight Committee

TPN	Total parenteral nutrition

UK	United Kingdom

### iii. TRIAL SUMMARY

| Trial Title | **D**oes **I**ntermittent **N**utrition **E**nterally **Normal**ise hormonal and metabolic responses to feeding in critically ill adults? The DINE-Normal proof of concept study. | |
| --- | --- | --- |
| Internal ref. no. (or short title) | **The DINE-Normal proof of concept study** | |
| Clinical Phase | Phase IIa | |
| Trial Design | Parallel group randomised open-label trial | |
| Trial Participants | 1. Adults (≥18) on intensive care 2. Gastric enteral nutrition (anticipated duration >48 hours) | |
| Planned Sample Size | 30 | |
| Treatment duration | Up to 48 hours | |
| Follow up duration | Until hospital discharge | |
| Planned Trial Period | 12 months | |
|  | Objectives | Outcome Measures |
| Primary | Compare and contrast the hormone and metabolite response to intermittent diurnal feeding in critically ill adults with the standard practice of continuous delivery. | Primary: peak plasma insulin and c-peptide level within 3 hours of bolus feed  Secondary: fatty acid, glycerol, triglyceride, urea, GLP-1, glucose, ketones |
| Secondary | Assess the clinical feasibility, tolerability, and efficacy of intermittent diurnal feeding in critically ill adults. | Feasibility: % target nutrition achieved, compliance with protocol  Tolerability: vomiting, aspiration, delayed gastric emptying, ileus, diarrhoea  Efficacy: ICU and hospital length of stay and mortality, delta-SOFA (day 0 – day 2) |
| Intervention | Intermittent diurnal gastric enteral nutrition | |
| Formulation, Dose, Route of Administration | Nutrison Protein Plus/Nutrison Concentrated via gastric tube at rate according to Intensive Care Unit Nutrition Guideline (2022) | |

### iv. FUNDING AND SUPPORT IN KIND

| **FUNDER(S)**  (Names and contact details of ALL organisations providing funding and/or support in kind for this trial) | **FINANCIAL AND NON FINANCIALSUPPORT GIVEN** |
| --- | --- |
| Southmead Hospital Charity Research Fund | Financial |
| Intensive Care Unit Southmead Hospital Research and Innovation Fund | Financial |

**v. ROLE OF TRIAL SPONSOR AND FUNDER**

North Bristol NHS Trust (NBT) acts as the trial sponsor under the Research Governance Framework for Health and Social Care. As Sponsor North Bristol NHS Trust has reviewed the protocol. The sponsor has responsibility for:

- Authorisation for clinical trials and research ethics committee opinion
- Good Clinical Practice and the conduct of clinical trial

Neither the Sponsor nor Funder (Southmead Hospital Charity Research Fund) are responsible for study design; data collection, analysis and interpretation; report writing or submission for publication.

**vi. ROLES AND RESPONSIBILITIES OF TRIAL MANAGEMENT COMMITEES**

Trial Management Group

A trial management group will be established to be responsible for day to day conduct of the study and overall supervision on behalf of the Sponsor and Funder and will ensure the trial is compliant with standards described in the Department of Health’s Research Governance Framework for Health and Social Care and the Guidelines for Good Clinical Practice. It will comprise study investigators with a Sponsor representative invited to all formal quarterly meetings. A subcommittee consisting of the Chief Investigator, Principal Investigator and clinical delivery staff will review study conduct and progress monthly.

Trial Oversight Committee

An oversight committee consisting of independent clinicians and one PPI representative will be established to review the study conduct and outcomes. It will receive reports from the quarterly meeting of the Trial Management Group.

**vii. PROTOCOL CONTRIBUTORS**

Dr Matt Thomas

Professor Tony Pickering

Dr Mike Ambler

Ms Danielle Milne

Dr Clodagh Beattie

Professor James Betts

Mr Harry Smith

Professor Javier Gonzalez

Professor Paul White

Dr Aravind Ramesh

Ms Borislava Borislavova

**viii. KEY WORDS:**

Critical illness; enteral nutrition; glycaemic control; insulin resistance; intensive care units, intermittent fasting; physiological stress

### ix. TRIAL FLOW CHART

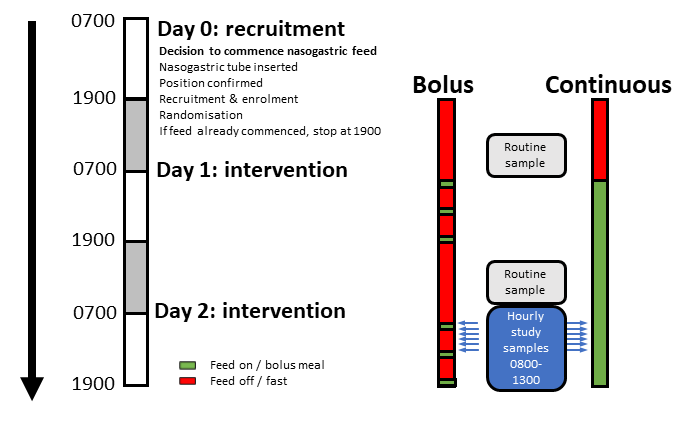

### 1 BACKGROUND

In the UK around 200,000 patients are admitted to critical care units annually ([ICNARC – Intensive Care National Audit & Research Centre](https://www.icnarc.org/); <https://www.sicsag.scot.nhs.uk/index.html>), approximately half of whom will be fed via a nasogastric (NG) tube (Binnekade et al., 2005). International guidelines emphasise the importance of providing early adequate enteral nutrition (Arabi et al., 2017; Singer et al., 2018; Compher et al., 2022) yet there remains surprisingly limited evidence to inform our approach.

There are several reasons to think that intermittent feeding with overnight fasting could be beneficial for critically ill patients rather than the usual continuous administration of NG feed. Evidence from animal and human studies indicates that aligning nutrition intake with the circadian rhythms of biological processes increases splanchnic blood flow and promotes the natural circadian pulsatile increases in ghrelin, insulin, and peptide YY levels (Chowdhury et al., 2016);  improves insulin sensitivity (Sjulin et al., 2020);  avoids the misalignment of circadian rhythms that is seen when food is taken in at night (Johnston, 2014; Skene et al., 2018); promotes regulated protein turnover; and modulates the immune system to promote tolerance to infection (Bear et al., 2018; Boutagouga Boudjadja et al., 2022; Ganeshan et al., 2019; Van Dyck & Casaer, 2019). All of these effects would improve tolerance to and recovery from critical illness.

Several studies of intermittent feeding have been conducted in the Intensive Care Unit (ICU). These have been small, each recruiting fewer than 200 patients and have failed to demonstrate improvements in outcomes (Bear et al., 2018; McNelly et al., 2020; Van Dyck & Casaer, 2019). Trial intermittent feeding regimens frequently continue to deliver food during the night or do not include a prolonged fasting period (Hiebert et al., 1981; Kadamani et al., 2014; MacLeod et al., 2007; McNelly et al., 2020; Nasiri et al., 2017; Rhoney et al., 2002; Serpa et al., 2003; Steevens et al., 2002). Primary outcomes have focused on measures of gastrointestinal tolerance while ignoring important hormonal, metabolic, circadian, and/or patient-centred outcomes, as highlighted in two recent reviews (Bear et al., 2018; Van Dyck & Casaer, 2019).

We (Smith, Gonzalez, & Betts) have recently completed a deep phenotyping study investigating the metabolic and immune consequences of intermittent versus continuous NG feed delivery in healthy volunteers (Figure below). This study represents one of the most detailed analyses of its kind and has highlighted a loss of the normal patterns of circulating glucose, fatty acid, triglycerides, and urea, as well as loss of the normal diurnal variation in insulin and glucagon-like peptide-1, alongside modulation of neutrophil metabolism, when feeding is delivered continuously.

These data, and ability to deliver these protocols and assays, puts our DINE-Normal trial team in a unique position, allowing direct ‘bench-to-bedside’ exploration of the interaction between critical illness and the mode of feed delivery, enabling comparison with the previous results from healthy volunteers. Complementing the team who carried out this work, is a group of clinicians with expertise and track record in the field of nutrition in the critically ill (Milne, Rooney), as well as wider clinical trial study design expertise (Pickering, Thomas) and animal studies of metabolism (Ambler, Pickering).

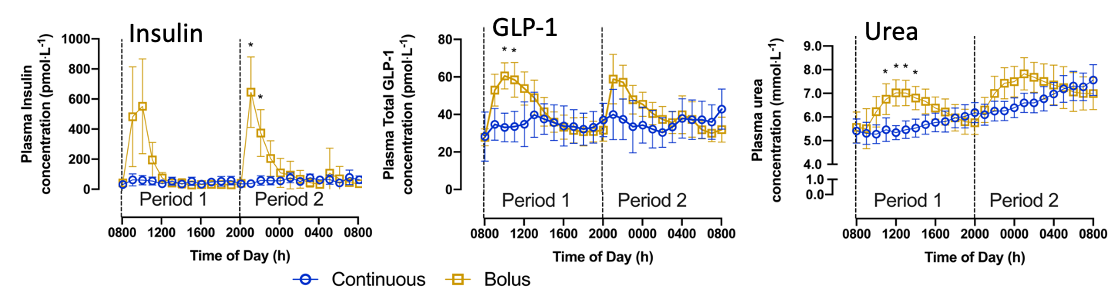

*Figure 1 Comparison of Bolus vs Continuous NG feed in healthy volunteers.  Blood profiles for hormones involved in metabolic control (insulin and GLP-1), and metabolites (urea) show marked post-prandial peaks in the bolus-fed subjects (n=8 per group). (Smith, Gonzalez & Betts, unpublished)*

**Aim**

The aim of this project is to establish whether intermittent feeding with overnight fasting, compared to the current standard of care in critically ill patients, produces the same improved physiological hormonal and metabolic responses to the meal challenge that are seen in healthy volunteers. If indeed we do see this effect in a sample of critically ill patients then the next step will be to use this data as the basis for either an MRC or NIHR-funded multicentre randomised controlled trial (RCT). This would further investigate the effects of continuous versus naturalistic feeding in this patient population and would include follow-up to establish any impact on patient-centred outcomes.

### 2 RATIONALE

Our recent study in healthy individuals has demonstrated clear disturbance of the normal circadian rhythms of metabolic, hormonal, and immune cell function when calories are delivered by continuous nasogastric infusion. It is likely that similar disruption to these key functions occurs in critically ill patients when fed continuously but this has not been fully investigated. This study will begin to address that knowledge gap.

Understanding the effects of continuous versus bolus feeding will help to optimise how nutrition is delivered to critically ill patients. This has the potential to provide several benefits: improved metabolic function with greater insulin sensitivity; reduced catabolism and sarcopenia, which would hasten rehabilitation and improve long-term functional status; altered immune response to improve outcomes in sepsis; better circadian rhythms with improved sleep/wake cycles, resulting in reduced delirium and less post-traumatic stress disorder.

The benefits for patients could include a shorter less complicated recovery from critical illness and lower mortality. For the NHS there are potential cost savings in ICU and hospital stay. Crucially implementation would not involve new drugs or technology and would be easy and cheap to do, potentially saving staff time.

The numbers involved are significant. Most recent (2021) summary statistics for England, Wales and Northern Ireland show 130,000 adult general ICU admissions with mortality of 24.2%. The average length of stay is more than 5 days in ICU and 19 in hospital with gastrointestinal support given on half of all patient days. It is rare to find an intervention that applies so broadly in critical care. Furthermore, the findings should be generalisable to less high-acuity patients in hospital and the community who receive nasogastric feeding. Finally, it might form the basis for a study of intermittent TPN delivery, extending the potential benefit and further reducing demand on NHS resource.

#### **2.1 Assessment and management of risk**

**2.1.1 Nasogastric tube insertion**

This is routine care in critically ill patients and presents no additional risk compared to standard practice. All insertions will be undertaken by trained professionals in line with local guidance. Both the nasal and oral routes are acceptable for the study.

**2.2.2 Enteral nutrition**

This is routine care in critically ill patients and presents no additional risk compared to standard practice. The Intensive Care Unit Nutrition Guidelines (2022) will be used in conjunction with this protocol and feed regimens will be reviewed by a specialist dietitian as standard in ICU. The usual enteral nutrition administration pumps and sets will be used.

**2.1.3 Bolus feed delivery**

The rate of bolus feed delivery for this study starts at 200 millilitres per hour (ml/hr). It is routine to return gastric aspirates of up to 250 millilitres (ml) in under 5 minutes in critically ill adults and common to administer 150-200 ml/hr for many hours in feed, water and medication. Gastric residual volumes are routinely monitored in all patients. Management of nausea, vomiting, diarrhoea, aspiration, ileus or constipation associated with enteral nutrition will be the same in both groups and will follow usual practice in the ICU as directed by the treating intensivist. Head up positioning to reduce the risk of regurgitation and aspiration is routine with the exception of patients with unstable spine or pelvic fractures. Bolus feeding presents no additional risk compared to standard practice.

**2.1.4 Hypo or hyperglycaemia**

Blood glucose levels will be managed according to North Bristol NHS Trust guidelines. Blood glucose is regularly monitored in critically ill adults. There is no additional risk compared to standard practice.

**2.1.5 Blood sampling**

Samples will be taken from existing routinely inserted arterial cannulas using aseptic non-touch technique. Additional samples for research will total 60 ml of blood over 48 hours which will not contribute significantly to the multifactorial anaemia that is usually seen in critically ill adults. There is no additional risk compared to standard practice.

### 3 OBJECTIVES AND OUTCOME MEASURES

**3.1**	**Primary objective**

Compare and contrast the hormone and metabolite response to intermittent diurnal feeding in critically ill adults with the standard practice of continuous delivery.

**3.2	Secondary objectives**

Assess the clinical feasibility, tolerability, and efficacy of intermittent diurnal feeding in critically ill adults.

**3.3	Primary outcome**

The primary outcome is peak plasma insulin (and c-peptide) within 3 hours of a bolus feed compared to the equivalent time period in continuous feed delivery.

**3.4	Secondary outcomes**

Secondary outcomes are:

- Endocrine and metabolic
  - Glucose
  - Ketones
  - Urea
  - GLP-1
  - Fatty acid
  - Triglyceride
  - Glycerol
- Feasibility
  - % target nutrition achieved (per 24 hour period)
  - Protocol compliance
- Tolerability
  - Episodes of vomiting/24 hour period
  - Episodes of aspiration of feed
  - Delayed gastric emptying (GRV>250ml x2 in a 24 hour period)
  - Ileus
  - Diarrhoea (passage of type 6 or 7 stool or >3 stool/24 hours)
- Efficacy
  - ICU and hospital length of stay
  - ICU and hospital mortality
  - Delta-SOFA (sequential organ failure assessment) between day 0 and day 2

**3.5	Table of outcomes**

| **Objectives** | **Outcome Measures** | **Timepoint(s) of evaluation** |
| --- | --- | --- |
| **Primary Objective**  Compare and contrast the hormone and metabolite response to intermittent diurnal feeding in critically ill adults with the standard practice of continuous delivery. | Primary: peak plasma insulin and c-peptide level within 3 hours of bolus feed  Secondary: fatty acid, glycerol, triglyceride, urea, GLP-1, glucose, ketones | See study flow chart for routine and research blood sampling time points |
| **Secondary Objectives**  Assess the clinical feasibility, tolerability, and efficacy of intermittent diurnal feeding in critically ill adults. | Feasibility: % target nutrition achieved, compliance with protocol  Tolerability: vomiting, aspiration, delayed gastric emptying, ileus, diarrhoea  Efficacy: ICU and hospital length of stay and mortality, delta-SOFA (day 0 – day 2) | Study intervention period  Study intervention and monitoring period  Day 2 (SOFA), at hospital discharge (length of stay, mortality) |

### 4 TRIAL DESIGN

A prospective, parallel group, randomised, open-label trial.

### 5 TRIAL SETTING

This single-centre study will be performed at Southmead Hospital, Bristol (North Bristol NHS Trust) in an adult general intensive care unit with 48 beds and 2500 admissions annually.

**6	PARTICIPANT ELIGIBILITY CRITERIA**

**6.1	Inclusion criteria**

- Adults (≥18) on intensive care
- Planned for gastric enteral nutrition (anticipated duration >48 hours)

**6.2	Exclusion criteria**

- >24 hours after commencement of enteral nutrition
- Gastrointestinal surgery or pathology
- Diabetic emergencies
- Pregnancy
- Parenteral or jejunal nutrition
- Trophic feed only (e.g. lactate >4)
- Prone positioning
- High risk of refeeding syndrome (NICE CG32)

### 7 TRIAL PROCEDURES

**7.1	Recruitment**

**7.1.1 Participant identification**

The usual care team will screen daily. Eligibility will be confirmed by health care professional on the delegation log and recorded in the ICU clinical information system (ICCA). This may be done remotely. A screening log will be kept.

**7.1.2	Screening**

No additional requirements above the clinical details needed for eligibility screening.

**7.2 Consent**

Informed consent will be sought from all participants. This is the responsibility of the Chief Investigator who may delegate this to persons who are trained and competent to participate according to the ethically approved protocol, principles of Good Clinical Practice (GCP) and Declaration of Helsinki.

Participants (or their Personal or Professional Consultee) are free to withdraw at any time from the study without giving reasons and without prejudicing any further treatment.

Many participants will lack capacity to consent at the time of enrolment as a consequence of illness and necessary medical interventions. As feeding is given to all patients, as early nutrition is given for maximum benefit, as prolonged feeding prior to intervention may bias the study, and as return of capacity is uncertain means it is not practicable to wait until capacity returns. The time critical nature of intervention and potential for significant additional distress for relatives in an emergency situation precludes seeking prior Personal Consultee opinion. As such an emergency waiver of consent model will be used, with informed consent sought once patients regain capacity. This approach was approved in our PPI group.

After confirmation of eligibility an emergency waiver form will be completed by the research team for patients entering the study prior to informed consent being obtained. Conformation of eligibility will be recorded in the patient medical notes and the Trial Site File.

Should an appropriate Personal Consultee be present at the time of confirmation of eligibility a brief verbal explanation of the study will be given by a member of the research team. If at this stage the Personal Consultee indicates the patient would not wish to participate in research no intervention will start and pseudonymised details will be entered in the screening log. If no such opinion is expressed the procedures described below will be followed.

The identified Personal Consultee will be given the Personal Consultee Information Sheet so they can inform the research team of the proposed participant’s wishes, feelings and values regarding being involved in the study. The Personal Consultee will be given adequate time to ask questions and to consider the patient’s participation in the study. If the Personal Consultee assessment of the person’s wishes, feelings and values indicate that the patient would want to participate in the research then the Personal Consultee will be asked to sign the Personal Consultee Declaration Form which will then be countersigned by a member of the research team. The Personal Consultee will retain a copy of the

signed Opinion Form. The original will be retained in the Trial Site File. The Opinion given by the Personal Consultee will be recorded in the medical notes.

Trial procedures will continue while a Personal Consultee is sought.

In the event that the patient is unable to give informed consent and no Personal Legal Representative is available or identifiable within 72 hours of admission to the Intensive Care Unit, a consultant unconnected with the study or an Independent Mental Capacity Advocate may act as a Professional Consultee. The Professional Consultee may also give an opinion for a patient to continue in the study if the Personal Consultee is undecided. The Professional Consultee will be informed about the trial by a member of the research team, given time to ask questions and asked to sign the Professional Consultee Declaration Form. The Professional Consultee will retain a copy of the signed Declaration Form. The original will be retained in the Trial Site File. Opinion from Professional Consultee will be recorded in the medical notes.

If a Personal Consultee is identified after a Professional Consultee has provided an opinion and the patient has not regained capacity the Personal Consultee will be provided with verbal and written information and their opinion sought as above. Personal consultee opinion, when given, will supersede the professional consultee opinion.

Trial procedures will continue while a Professional Consultee is sought.

Patients for whom an opinion is given by a Personal Consultee or Professional Consultee will be informed of their study participation by a member of the research team once they have regained the capacity to understand the trial details providing this is before discharge from acute hospital. The research team will discuss the study with the patient and the patient will be given a copy of the Patient Information Sheet (PIS). The patient will be given time to read the information sheet, to ask questions and to consider their participation in the study. The patient will be asked for consent to continue their participation in the study and asked to sign the Informed Consent Form. The patient will retain one copy of the signed Consent Form. Another copy will be placed in the patient’s medical records whilst the original will be retained in the Trial Site File.

**7.3	Randomisation**

Allocation 1:1 to intervention or control stratified by sex.

**7.3.1	Method of randomisation**

The National Cancer Institute Clinical Trial Randomization Tool [Clinical Trial Randomization Tool - Clinical Trial Randomization Tool (cancer.gov)](https://ctrandomization.cancer.gov/tool/)) was used to generate the allocation sequence which was concealed using sealed opaque envelopes stored in a locked cabinet in the Intensive Care Unit.

**7.4	Baseline data**

Baseline data will be collected as follows:

- Sex
- Ethnicity
- Age
- Weight
- Height
- Severity of illness (APACHE II and ICNARC physiology score)
- Organ failure assessment (Sequential Organ Failure Assessment)
- Primary reason for admission to ICU
- Time of admission
- Time of gastric tube insertion
- Time enteral feeding started prior to enrolment
- Time of enrolment
- Presence/absence of diabetes (including type if present)
- Medication (insulin, oral hypoglycaemic agents, statins)

**7.5	Trial assessments**

| Assessment | Measurement | Timepoint(s) of evaluation |
| --- | --- | --- |
| Hormonal and metabolic | Insulin, C-peptide, fatty acid, glycerol, triglyceride, GLP-1, glucose, ketones urea | During study intervention period (see Section 8.1) |
| Acid-base balance | Arterial blood gas (pH, bicarbonate, lactate, base excess) | With routine daily blood sampling |
| Haematology | Haematocrit, haemoglobin, platelet count, red cell distribution width, red blood cell count, white blood cell count, neutrophils, lymphocytes.  Activated partial thromboplastin time, prothrombin time, International Normalised Ratio (INR)  C-reactive protein | With routine daily blood sampling |
| Biochemistry | Albumin, alkaline phosphatase, alanine aminotransferase, calcium, creatinine, magnesium, phosphate, potassium, sodium, total bilirubin, urea, creatinine, eGFR | With routine daily blood sampling |
| Clinical | SOFA  Mortality and length of stay | Baseline, day 2  ICU and hospital discharge |
| Nutrition | Vomiting  Diarrhoea  Delayed gastric emptying  Ileus  Aspiration  Total enteral nutritional intake | During study intervention period |

**7.6	Stopping criteria**

The trial intervention will be terminated if any of the following events occur:

- Death
- Discharge from the ICU
- Withdrawal of life sustaining treatment
- Withdrawal of consent/request to withdraw from study by patient (or personal or professional consultee if patient unable to consent)
- Decision by attending clinician that the delivery of feed should be stopped or adjusted on safety grounds.
- Development of any of the conditions listed in the exclusion criteria.

**7.7	Participant withdrawal**

Participants, or their personal or professional consultees, may withdraw from the study at any time. No reason need be given and usual medical care will not be affected. Only data essential for study monitoring and oversight will be retained.

**7.8	Storage and analysis of clinical samples**

Samples will be immediately distributed into tubes coated in ethylenediaminetetraacetic acid (EDTA) as well as a serum tube containing polystyrene beads. All samples will be centrifuged at 3461 x g for 10 minutes for removal of the plasma or serum supernatant then frozen at -80°C pending analysis.

Analysis of samples at the University of Bath will use standard techniques and commercially available assay kits for the relevant primary and secondary outcomes.

Additional blood samples will not be stored for the purposes of the study.

The blood samples will not be stored beyond analysis of the primary and secondary outcomes.

#### **7.9 End of trial**

The trial will end upon completion of the follow up period for the final participant or if required by the Sponsor, Research Ethics Committee or the Trial Oversight Committee.

### 8 TRIAL INTERVENTION

**8.1	Name and description of trial intervention**

The intervention is an adjustment to the pattern of delivery of gastric feed. The feed type will follow the Intensive Care Unit Nutrition Guideline (2022).

The intervention period consists of 48 hours from the initiation of the first overnight fast (1900 on day 0) to the establishment of the usual ICU Nutrition guideline enteral feeding regimen (1900 on day 2). There will be a further 12 hours of monitoring for adverse events potentially attributable to intervention.

For intermittent diurnal feeding bolus feeds will start at 0800, 1300 and 1800 and be given over 30 to 60 minutes. On study day 1 each feed will be 200ml. On study day 2 each feed will be 1/3 of total required volume of feed based on hypocaloric targets for days 3-6 as in the Intensive Care Unit Nutrition Guideline (2022).

Continuous feeding will start at 0800 on study day 1 at 25ml/hr. At 0800 on study day 2 the rate of continuous feeding will be adjusted to meet the hypocaloric targets for days 3-6 as in the Intensive Care Unit Nutrition Guideline (2022).

Reduced feed volumes will be used in patients who are fluid restricted or hyperkalaemic in the absence of renal replacement therapy as per the Intensive Care Unit Nutrition Guideline (2022).

The Intensive Care Unit Nutrition Guideline (2022) gives the basis for study management of gastric residual volumes. During the intervention period GRV will be checked in both groups at 0800, 1300 and 1800 with further checks at 2200, 0200 and 0600.

Research blood samples (10ml per sample) will be taken the day after recruitment to the study at 0800 (before the start of feed in the intermittent group), 0900 (at the end of feed in the intermittent group), 1000, 1100, 1200 and 1300 (before start of feed in the intermittent group).

**8.2	Regulatory status of enteral nutrition**

The products used for enteral feeding have Advisory Committee on Borderline Substances (ACBS) approval.

**8.3	Product Characteristics**

Data cards (Nutrison Protein Plus and Nutrison Concentrated) are shown in the appendix.

**8.4	Storage and supply**

As enteral feed is stocked routinely storage and supply will follow usual practice.

**8.5	Dosage schedules**

Patients in the trial will be randomised to receive either intermittent diurnal or continuous feed delivery for 48 hours (see s8.1). Prescribed targets will follow the Intensive Care Unit Nutrition Guideline (2022).

If feed is interrupted or delayed (for example in the case of transfer for imaging or surgery) the following adjustments to the feeding regimen will be made:

- Intermittent diurnal feed: if there is two hours or more until the next scheduled feed the missed feed will be given in entirety followed by the next feed at the scheduled time. If there is less than two hours until the next scheduled feed the missed feed will be given in entirety followed by a one hour gap and then the next scheduled feed.
- Continuous feed: the feed rate will be increased to compensate for the hours missed to achieve the prescribed 24 hour target.

**8.6	Dosage modifications**

The feeding regimen will be reviewed at meetings of the Trial Management Group.

**8.7	Known drug reactions and interaction with other therapies**

Enteral feed may interact with the absorption of some medications including certain antiretrovirals and antibiotics, antiepileptics and immunosuppressants. Daily medication review by a Pharmacist is routine in the ICU. Alternative routes of administration and therapeutic drug monitoring will be undertaken as advised.

#### **8.8 Concomitant medication**

There is no restriction on concomitant medication.

**8.****9	Assessment of compliance**

Compliance with the intervention will be assessed by the use of routinely collected patient data on ICU, namely the feeding administration record for each patient available in ICCA.

### 9 SAFETY AND MONITORING

**9.1	Definitions**

| **Term** | **Definition** |
| --- | --- |
| **Adverse Event (AE)** | Any untoward medical occurrence in a participant to whom a medicinal product has been administered, including occurrences which are not necessarily caused by or related to that product. |
| **Adverse Reaction (AR)** | An untoward and unintended response in a participant to an investigational medicinal product which is related to any dose administered to that participant.  The phrase "response to an investigational medicinal product" means that a causal relationship between a trial medication and an AE is at least a reasonable possibility, i.e. the relationship cannot be ruled out.  All cases judged by either the reporting medically qualified professional or the Sponsor as having a reasonable suspected causal relationship to the trial medication qualify as adverse reactions. It is important to note that this is entirely separate to the known side effects listed in the SmPC. It is specifically a temporal relationship between taking the drug, the half-life, and the time of the event or any valid alternative aetiology that would explain the event. |
| **Serious Adverse Event (SAE)** | A serious adverse event is any untoward medical occurrence that:   - results in death - is life-threatening - requires inpatient hospitalisation or prolongation of existing hospitalisation - results in persistent or significant disability/incapacity - consists of a congenital anomaly or birth defect   Other ‘important medical events’ may also be considered serious if they jeopardise the participant or require an intervention to prevent one of the above consequences.  NOTE: The term "life-threatening" in the definition of "serious" refers to an event in which the participant was at risk of death at the time of the event; it does not refer to an event which hypothetically might have caused death if it were more severe. |
| **Serious Adverse Reaction (SAR)** | An adverse event that is both serious and, in the opinion of the reporting Investigator, believed with reasonable probability to be due to one of the trial treatments, based on the information provided. |
| **Suspected Unexpected Serious Adverse Reaction (SUSAR)** | A serious adverse reaction, the nature and severity of which is not consistent with the information about the medicinal product in question set out in the reference safety information:   - in the case of a product with a marketing authorisation, this could be in the summary of product characteristics (SmPC) for that product, so long as it is being used within its licence. If it is being used off label an assessment of the SmPCs suitability will need to be undertaken. - in the case of any other investigational medicinal product, in the investigator’s brochure (IB) relating to the trial in question |

**9.2	Operational definitions for (S)AEs**

The patient group of interest are extremely medically unwell and as such adverse events are common. All events will be considered in the context of the patient’s individual clinical status and a clinical decision made by the medically qualified clinicians on the delegation log as to the likely causality. The AE monitoring period extends for 12 hours after the end of the intervention period. Where AE are secondary outcomes of the study there is no need for additional safety reporting.

**9.3	Recording and reporting of SAEs, SARs AND SUSARs**

Adverse events will be recorded and reported in accordance with North Bristol NHS Trust’s Safety Reporting SOP (SOP 013c - Safety Reporting - non CTIMP).

All SAEs occurring during the intervention monitoring period must be recorded and emailed to the CI within 24 hours of the research team becoming aware of the event.

For each SAE the following information will be collected:

- full details in medical terms and case description
- event duration (start and end dates, if applicable)
- action taken
- outcome
- seriousness criteria
- causality (i.e. relatedness to trial drug / investigation), in the opinion of the investigator
- whether the event would be considered anticipated.

Any change of condition or other follow-up information should be emailed to the CI as soon as it is available or at least within 24 hours of the information becoming available. Events will be followed up until the event has resolved or a final outcome has been reached.

All SAEs assigned by the PI or delegate (or following central review) as both suspected to be related to the study intervention and unexpected will be classified as SUSARs. The CI will inform the sponsor, the REC and Marketing Authorisation Holder of SUSARs within the required expedited reporting timescales.

Events that represent the natural history of critical illness and would be expected in patients undergoing intensive care treatment, should NOT be reported. These include but are not limited to death, persistent or significant disability or incapacity, acute organ failure and hospital acquired infection.

**9.4	Responsibilities**

**9.4.1. Principal Investigator (PI)**

Checking for AEs and ARs during the study period.

1. Using medical judgement in assigning seriousness, causality and whether the event/reaction was anticipated using the Reference Safety Information approved for the trial.
2. Using medical judgement in assigning seriousness and causality and providing an opinion on whether the event/reaction was anticipated using the Reference Safety Information approved for the trial.
3. Ensuring that all SAEs are recorded and reported to the sponsor within 24 hours of becoming aware of the event and provide further follow-up information as soon as available. Ensuring that SAEs are chased with Sponsor if a record of receipt is not received within 2 working days of initial reporting.
4. Ensuring that AEs and ARs are recorded and reported to the sponsor in line with the requirements of the protocol.

**9.4.2. Chief Investigator (CI) or delegate**

1. Clinical oversight of the safety of patients participating in the trial, including an ongoing review of the risk / benefit.
2. Using medical judgement in assigning the SAEs seriousness, causality and whether the event was anticipated (in line with the Reference Safety Information) where it has not been possible to obtain local medical assessment.
3. Using medical judgement in assigning whether and event/reaction was anticipated or expectedness in line with the Reference Safety Information.
4. Immediate review of all SUSARs.
5. Review of specific SAEs and SARs in accordance with the trial risk assessment and protocol as detailed in the Trial Monitoring Plan.
6. Assigning Medical Dictionary for Regulatory Activities (MedDRA) or Body System coding to all SAEs and SARs.

**9.4.3 Sponsor**

1. Central data collection and verification of AEs, ARs, SAEs, SARs and SUSARs according to the trial protocol onto a database.
2. Reporting safety information to the CI, delegate or independent clinical reviewer for the ongoing assessment of the risk / benefit according to the Trial Monitoring Plan.
3. Reporting safety information to the independent oversight committees identified for the trial according to the Trial Monitoring Plan.
4. Expedited reporting of SUSARs to the Competent Authority (HRA)
5. and REC within required timelines.
6. Notifying Investigators of SUSARs that occur within the trial.
7. Checking for (annually) and notifying PIs of updates to the Reference Safety Information for the trial.
8. Preparing standard tables and other relevant information for the DSUR in collaboration with the CI and ensuring timely submission to the HRA and REC.

##### 9.5 Notification of deaths

Only deaths that are assessed to be caused by the study intervention will be reported to the sponsor. This report will be immediate.

##### 9.6 Reporting urgent safety measures

If any urgent safety measures are taken the CI/Sponsor shall immediately and in any event no later than 3 days from the date the measures are taken, give written notice to the HRA and the relevant REC of the measures taken and the circumstances giving rise to those measures.

#### **9.7 The type and duration of the follow-up of participants after adverse reactions.**

Adverse events and reactions will be recorded and reported up to 12 hours after the cessation of the study intervention. Follow-up will be coordinated with the admitting medical team. Any SUSAR will need to be reported to the Sponsor irrespective of how long after the intervention period the reaction has occurred until resolved.

### 10 STATISTICS AND DATA ANALYSIS

**10.1	Sample size calculation**

A sample size of 30 participants will be used.

In data from healthy subjects the intermittent group had a mean peak plasma insulin concentration at 2 hours after the bolus of 373±204 pmol/l (mean ±SD) compared to the continuous feed group 58±41 pmol/l. This effect size of 2.14 would require 6 subjects per group to have a 90% power (p=0.05). A smaller and/or more variable response in critically ill patients would be clinically relevant.

The sample size calculation used a conservative effect size based on this healthy volunteer data from our group and was adjusted following advice from Dr Paul White. The lower limit of the 80% confidence interval for effect size was used in the updated calculation to reflect the small sample size in the volunteer data and the possibility the assumption of equal variances does not hold. With a lower limit of 1.26 the study would have 80% power with 11 patients per group, and 90% power with 15 patients per group.

The overall sample size has been adjusted to 30 to allow for this more conservative effect size estimate while retaining at least 80% power in the case of more than 25% drop out.

**10.2	Planned recruitment rate**

Seventy-five patients receive gastric feeding on the ICU per month at Southmead Hospital. A maximum of one patient per day will be recruited, in order to facilitate timely sample collection.

**10.3	Statistical analysis plan**

Analysis will be undertaken by the study statistician (Dr Paul White) blinded to group allocation. Normality will be tested and appropriate parametric or non-parametric tests used for the primary outcome. Additional analysis of the insulin/c-peptide response (e.g. area under the curve) and of secondary outcomes will be undertaken as appropriate or as directed by trial committees. A p value < 0.05 will be taken as significant.

**10.3.1	Summary of baseline data and flow of patients**

Baseline demographic and clinical variables will be summarised. Descriptive summaries of the distributions of continuous baseline variables will be presented in terms of percentiles, while discrete variables are summarised in terms of frequencies and percentages. No formal statistical testing will be undertaken for differences in baseline characteristics.

**10.3.2	Primary outcome analysis**

Mixed ANOVA will be used for analysis of peak plasma insulin/c-peptide.

**10.3.3	Secondary outcome analysis**

Secondary physiological outcomes (plasma fatty acid, glycerol, triglyceride, urea, GLP-1, blood glucose, and ketones) will also be analysed using mixed ANOVA.

Other secondary outcomes will be reported using descriptive statistics.

**10.4	Interim and subgroup analysis**

No interim or subgroup analysis is planned.

**10.5	Participant population**

All participants will be analysed as randomised (intention-to-treat analysis).

**10.6	Procedure(s) to account for missing or spurious data**

Data will be recorded as missing if queries are unable to recover missing data. Missing data will not be imputed for analysis.

### 11 DATA MANAGEMENT

#### **11.1 Data collection tools and source document identification**

Each participant will be assigned a unique study number, allocated at enrolment, for use on CRFs and other trial documents and samples. The fact that the patient is participating in a clinical trial will be added to the patient’s medical record contemporaneously, along with the date of consultee opinion, and patient informed consent when capacity is regained.

Data will be collected on a paper CRF. Data recorded in the CRF will be consistent and verifiable with source data in source documents other than the CRF (e.g. medical record, laboratory reports, nurses’ notes and participant questionnaires). Appropriate medical and research records will be maintained for this study, in compliance with ICH E6 GCP, Section 4.9 and regulatory and institutional requirements for the protection of confidentiality of subjects.

All paper forms shall be filled in using black ballpoint pen. All data requested on the CRF must be recorded. All missing data must be explained. If a space on the CRF is left blank because the procedure was not done or the question was not asked, write “N/D”. If the item is not applicable to the individual case, write “N/A”. Errors shall be lined out but not obliterated and the correction inserted, initialled and dated.

Paper copies of the CRF will be kept in a secure location (locked cabinet).

An online Redcap database will be used to store and assimilate clinical and assay data. The database will protect patient information in line with (i) the Data Protection Act 1998 until 24 May 2018, and (ii) the General Data Protection Regulation, as from time to time amended from 25 May 2018. Trial staff will ensure that the participants’ confidentiality is maintained through protective and secure handling and storage of patient information at the trial centres (as relevant). All documents will be stored securely and only accessible by trial staff and authorised personnel. Data will be collected and retained in accordance with the relevant data protection legislation. The North Bristol NHS Trust SOP on Data Management will be followed (SOP 017 - Essential Documents).

Personal identifiers will be held only until the results are available in order to offer a lay summary to participants and families.

#### **11.2 Access to Data**

Direct access will be granted to authorised representatives from the Sponsor, host institution and the regulatory authorities to permit trial-related monitoring, audits and inspections- in line with participant consent.

11.3 Archiving

Archiving will be authorised by the Sponsor following submission of the end of trial report. All essential documents will be archived for a minimum of 5 years after completion of trial. Destruction of essential documents will require authorisation from the Sponsor. North Bristol NHS Trust SOP on Archiving (SOP 010 - Archiving) will be followed.

##### 12 MONITORING, AUDIT & INSPECTION

The study will be monitored in accordance with North Bristol NHS Trust’s Monitoring SOP (SOP 014 Monitoring). All trial related documents will be made available on request for monitoring and audit by North Bristol NHS Trust, the Research Ethics Committee and for inspection by the Medicines and Healthcare products Regulatory Authority or other licensed bodies. The monitoring plan will be developed and agreed by the sponsor.

### 13 ETHICAL AND REGULATORY CONSIDERATIONS

13.1 **Research Ethics Committee (REC) review & reports**
- Before the start of the trial, approval will be sought from a REC and the HRA, as well as confirmation of capacity and capability from the sponsor prior to starting, for the trial protocol, informed consent forms and other relevant documents e.g. advertisements and GP information letters.
- Substantial amendments that require review by REC will not be implemented until the REC grants a favourable opinion for the trial.
- All correspondence with the REC will be retained in the Trial Site File.
- An annual progress report (APR) will be submitted to the REC within 30 days of the anniversary date on which the favourable opinion was given, and annually until the trial is declared ended. It is the Chief Investigator’s responsibility to produce the annual reports as required.
- The Chief Investigator will notify the REC of the end of the trial. If the trial is ended prematurely, the Chief Investigator will notify the REC, including the reasons for the premature termination.
- Within one year after the end of the trial, the Chief Investigator will submit a final report with the results, including any publications/abstracts, to the REC

**13.2	Peer review**

The study summary has been reviewed by the funding panel, including lay members, and by the research advisory group. The protocol has been reviewed by Sponsor, research advisory group and the investigators.

**13.3	Public and Patient Involvement**

The study summary has been reviewed by the lay panel of the Funder. The protocol and patient facing documentation has been reviewed by the study patient advisory group.

**13.4	Regulatory Compliance**

The study will be performed subject to favourable opinion/authorisation/permission from all necessary regulatory and other bodies. This includes but is not limited to REC, HRA, NHS Trusts.

This study will be conducted in accordance with:

- International Conference for Harmonisation of Good Clinical Practice (ICH GCP) guidelines
- Research Governance Framework for Health and Social Care

**13.5	Protocol compliance**

Prospective, planned deviations or waivers to the protocol are not allowed under the UK regulations on Clinical Trials and must not be used e.g. it is not acceptable to enrol a participant if they do not meet the eligibility criteria or restrictions specified in the trial protocol

Accidental protocol deviations can happen at any time. They must be adequately documented on the relevant forms and reported to the Chief Investigator and Sponsor immediately. Deviations from the protocol which are found to frequently recur are not acceptable, will require immediate action and could potentially be classified as a serious breach.

The North Bristol NHS Trust SOPs ‘Identifying and Preventing Non-compliance with Good Clinical Practice or the Protocol’ (SOP 012b - Identifying and Preventing Non-Compliance with Good Clinical Practice or the Protocol) and ‘Managing Breaches of Good Clinical Practice or the Protocol’ (SOP 012- Managing Breaches of GCP or the Protocol) will be followed. The definition of non-compliance given in the former document will be used for the study.

##### 13.6 Notification of Serious Breaches to GCP and/or the protocol

A “serious breach” is a breach which is likely to effect to a significant degree:

a. the safety or physical or mental integrity of the participants of the trial; or
b. the scientific value of the trial

The sponsor will be notified immediately of any case where the above definition applies during the trial conduct phase. The sponsor will notify the licensing authority in writing of any serious breach of:

a. the conditions and principles of GCP in connection with that trial; or
b. the protocol relating to that trial, as amended from time to time, within 7 days of becoming aware of that breach

The North Bristol NHS Trust SOP for Managing Breaches of Good Clinical Practice or Protocol will be followed.

**13.7	Data protection and patient confidentiality**

All investigators and trial site staff will comply with the requirements of the General Data Protection Regulation with regards to the collection, storage, processing and disclosure of personal information.

Participants will be assigned a unique study number to be used throughout their participation in the trial. Any personal data recorded will be regarded as confidential, and any information that would allow individual participants to be identified will not be released into the public domain.

The data and the linking code will be stored in separate locations using encrypted files within password protected folders. The Chief Investigator will be the data custodian.

13.8	Conflicts of interest

None of the research team, investigator teams, and the sponsor has any financial or other conflict of interest. All members of the oversight committees will declare any potential conflicts of interest as part of their membership agreement.

13.9	Indemnity

This is an NHS-sponsored research study. For NHS sponsored research HSG(96)48 reference no.2 refers. If there is negligent harm during the clinical trial when the NHS body owes a duty of care to the person harmed, NHS indemnity covers NHS staff, medical academic staff with honorary contracts, and those conducting the trial. NHS indemnity does not offer no-fault compensation and is unable to agree in advance to pay compensation for non-negligent harm.

13.10	Amendments

Potential amendments will be discussed by the Chief Investigator (advised by the Trial Management Group), the trial manager and Sponsor. North Bristol NHS Trust SOP on Research Study Amendments (SOP 003 - Research Study Amendments) will be followed in this study.

Under the Medicines for Human Use (Clinical Trials) Regulations 2004, the sponsor may make a non-substantial amendment at any time during a trial. If the sponsor wishes to make a substantial amendment to the REC application or the supporting documents, the sponsor must submit a valid notice of amendment to the REC for consideration. The REC will provide a response regarding the amendment within 35 days of receipt of the notice. It is the sponsor’s responsibility to decide whether an amendment is substantial or non-substantial for the purposes of submission to the REC.

Amendments will also be notified to the national coordinating function of England and communicated to the participating organisations (NHS R&D office and local research team) departments of participating sites to assess whether the amendment affects the NHS permission for that site. Note that some amendments that may be considered to be non-substantial for the purposes of REC still need to be notified to NHS R&D (e.g. a change to the funding arrangements).

**13.11	Post trial care**

After the trial intervention period participants will follow the Intensive Care Unit Nutrition Guideline (2022).

**13.12	Access to the final trial dataset**

The co-applicants, collaborators and Sponsor will have access to the final trial dataset. Applications for access to the final trial dataset will be considered by the Chief Investigator and Sponsor after publication of trial results.

##### 14 DISSEMINATION POLICY

##### 14.1 Dissemination policy

The trial will be reported in accordance with the Consolidated Standards of Reporting Trials (CONSORT) guidelines ([www.consort-statement.org](http://www.consort-statement.org)). The main report will be written by the Trial Management Group with authorship determined according to the internationally agreed criteria for authorship ([www.icmje.org](http://www.icmje.org)).

The report will be presented at scientific and clinical meetings and uploaded to a pre-print server prior to publication in an open access peer-reviewed journal. Participants will be asked if they wish to have a lay summary of the findings.

##### 16. APPENDICES

**16.1	Risk**

| Risks associated with trial interventions  ☒ A ≡ Comparable to the risk of standard medical care  ☐ B ≡ Somewhat higher than the risk of standard medical care  ☐ C ≡ Markedly higher than the risk of standard medical care | | | |
| --- | --- | --- | --- |
| Justification:  The risk associated with the trial intervention is comparable to the risk of standard medical care as it represents a more naturalistic pattern for the delivery of feed. This feed is already widely used in clinical practice, has local formulary approval, and will be given under the same dietician/nutritionist guidance and oversight as any other enteral feeding in these patients. | | | |
| What are the key risks related to therapeutic interventions you plan to monitor in this trial? | | How will these risks be minimised? | |
| Intervention | Body system/Hazard | Activity | Frequency |
| Nasogastric feed delivered in 3 equal divided boluses | See section 2.1 | - Level 3 or level 2 care - Routine monitoring of gastrointestinal function & blood glucose | - Continuous - See section 7.7 |
| Outline any other processes that have been put in place to mitigate risks to participant safety (e.g. DMC, independent data review, etc.)  See sections 2.1 and 9.4 | | | |

**16.2	Intensive Care Unit Nutrition Guideline (2022)**

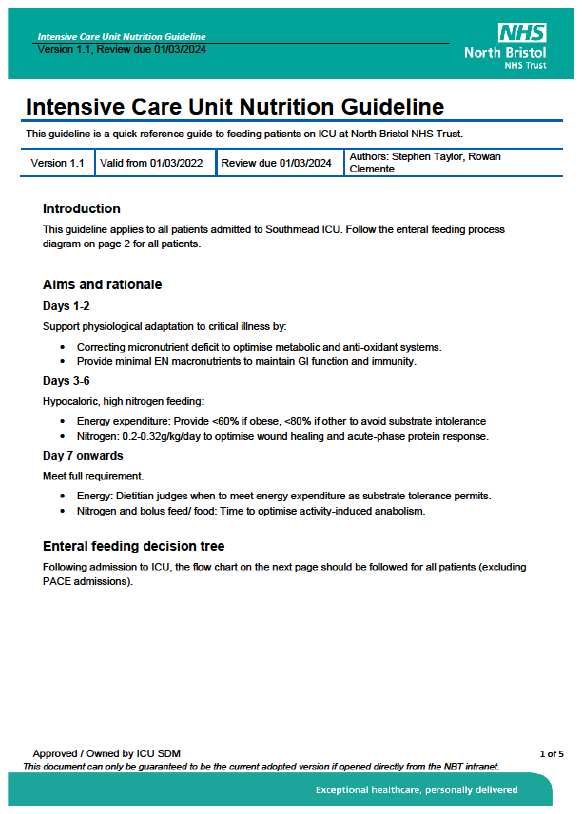

**16.3	Management of blood glucose**

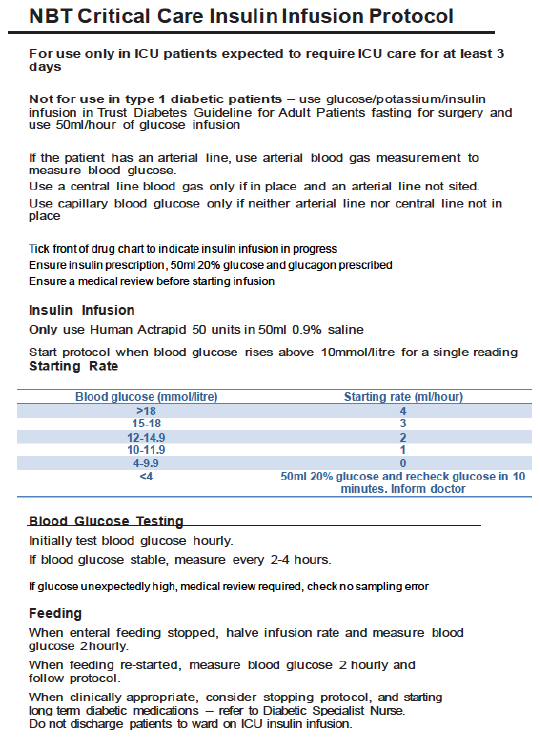

**16.4**	**Schedule of Procedures**

| **Procedures** | **Visits (insert visit numbers as appropriate)** | | | |
| --- | --- | --- | --- | --- |
|  | **Screening** | **Baseline** | **Treatment Phase (see schedule above)** | **Follow Up** |
| Informed consent (see details above) |  | 2 | 3 | 4 |
| Demographics | 1 |  |  |  |
| Medical history | 1 |  |  |  |
| Physical examination |  | 2 |  |  |
| Vital signs | 1 |  | 3 |  |
| Concomitant medications |  | 2 |  |  |
| ECG |  | 2 |  |  |
| Laboratory tests |  | 2 | 3 |  |
| Eligibility assessment | 1 |  |  |  |
| Compliance |  |  | 3 |  |
| Assessment 1 (blood tests for insulin / c-peptide levels) |  |  | 3 |  |
| Assessment 2 plasma fatty acid, glycerol, triglyceride, urea, GLP-1, blood glucose, & ketones;) |  |  | 3 |  |
| Assessment 3 (clinical assessment of gastrointestinal function and calorie delivery) |  |  | 3 |  |
| Adverse event assessments |  |  | 3 | 4 |
| Physician’s Withdrawal Checklist |  |  | 3 |  |

**16.5	Data card Nutrison Protein Plus**

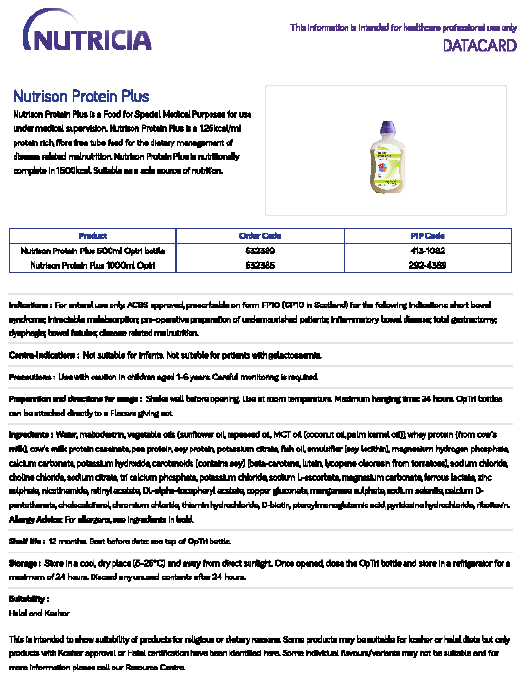

**16.6	Data card Nutrison Concentrated**

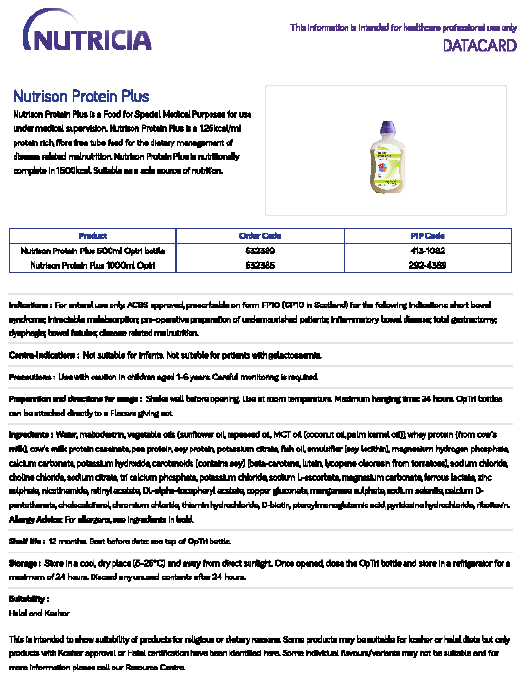

**16.7**	**Amendment History**

| **Amendment No.** | **Protocol version no.** | **Date issued** | **Author(s) of changes** | **Details of changes made** |
| --- | --- | --- | --- | --- |

List details of all protocol amendments here whenever a new version of the protocol is produced.

Protocol amendments must be submitted to the Sponsor for approval prior to submission to the REC committee or HRA.

**16.8	Investigators and contributions**

Dr Matt Thomas	Chief Investigator

Professor Tony Pickering	Conception, study design and interpretation

Dr Mike Ambler	Study design and interpretation

Ms Danielle Milne	Study design and interpretation, specialist dietetic	advice

Dr Clodagh Beattie	Principal Investigator, study delivery

Professor James Betts	Conception, study design and interpretation

Mr Harry Smith	Conception, study design and interpretation, laboratory	analysis

Professor Javier Gonzalez	Study design and interpretation

Professor Paul White	Study statistician

Dr Aravind Ramesh	Study design and interpretation

Ms Borislava Borislavova	Study design and delivery

**16.9	North Bristol NHS Trust Standard Operating Procedures**

All North Bristol NHS Trust SOPs are available on request from.
